# Multi-state network meta-analysis of progression and survival data

**DOI:** 10.1101/2020.11.13.20231332

**Authors:** Jeroen P. Jansen, Devin Incerti, Thomas A. Trikalinos

## Abstract

Multiple randomized controlled trials, each comparing a subset of competing interventions, can be synthesized by means of a network meta-analysis to estimate relative treatment effects between all interventions in the evidence base. Here we focus on estimating relative treatment effects for time-to-event outcomes. Cancer treatment effectiveness is frequently quantified by analyzing overall survival (OS) and progression-free survival (PFS). We introduce a method for the joint network meta-analysis of PFS and OS that is based on a time-inhomogeneous tri-state (stable, progression, and death) Markov model where time-varying transition rates and relative treatment effects are modeled with parametric survival functions or fractional polynomials. The data needed to run these analyses can be extracted directly from published survival curves. We demonstrate use by applying the methodology to a network of trials for the treatment of non-small-cell lung cancer. The proposed approach allows the joint synthesis of OS and PFS, relaxes the proportional hazards assumption, extends to a network of more than two treatments, and simplifies the parameterization of decision and cost-effectiveness analyses.

## 1 Introduction

Randomized controlled trials (RCTs) are considered the most appropriate study design to obtain evidence regarding relative treatment effects. However, an individual RCT rarely includes all alternative interventions of interest, and as such does not provide all the information needed to select the best alternative. Typically, the evidence base consists of multiple RCTs, each of which compares a subset of the interventions of interest. If each of these trials has at least one intervention in common with another trial such that the evidence base is represented by a single connected network, a network meta-analysis (NMA) can estimate relative treatment effects between all the competing interventions in the evidence base^1^.

Often there is an interest in estimating the relative treatment effects of alternative interventions regarding time-to-event outcomes. For example, in oncology treatment efficacy is often quantified by analyzing time from treatment initiation to the occurrence of a particular event. Very commonly, studies report data on overall survival (OS), where the event is death from any cause, and on progression-free survival (PFS), where the event is death from any cause or disease progression, whichever occurred first^2^.

NMA of time-to-event outcomes with a single effect measure per study are based on the proportion of patients alive at a specific time point, median survival, or reported hazard ratio (HR)^3^. The limitation of a NMA of survival at a specific time point is that we only focus on the cumulative effect of treatment at that time point and ignore the variation in effects over time up to, as well as beyond, that time point. NMAs of median survival times have similar limitations. The HR summarizes the treatment effect for the complete follow-up period of the trials, but only represents the treatment effect for each time point if the proportional hazards (PH) assumption holds. If the PH assumption is violated, trial specific HRs represent an average effect over the follow-up period, which can cause biased estimates in a NMA if trials have different lengths of follow-up.

As an alternative to a NMA with a univariate treatment effect measure, we can also use a multivariate treatment effect measure that describes how the relative treatment effects change over time^3^. Ouwens et al., Jansen, and Cope et al presented methods for NMA of time-to-event outcomes where the hazard functions of the interventions in a trial are modeled using parametric survival functions or fractional polynomials and the difference in the parameters are considered the multi-dimensional treatment effects, which are synthesized across studies^4–7^. By incorporating time-related parameters, these NMA models can be fitted more closely to the available data.

Both PFS and OS of an intervention determine its value and can inform decision-making. In combination with a baseline survival function for a reference treatment, the multivariate NMA models embedded in parametric survival functions can form the basis for partitioned-survival cost-effectiveness models^8^. Frequently, the pooled PFS and OS curves need to be extrapolated over time in order to obtain estimates of the expected quality adjusted life-years before and after disease progression. Since the separate meta-analyses of PFS and of OS data ignore the correlation between the outcomes, any required extrapolation may result in possible crossing of PFS and OS curves. A state-transition model with three health states – stable (pre-progression), progression, and death – with parametric hazard functions for the three corresponding transitions avoids this issue. If we have individual patient data (IPD) regarding time to progression, time to death, and censoring for all trials included in the NMA, we can estimate these hazard functions using a statistical model with the same tri-state structure and avoid any inconsistency between the clinical evidence synthesis and the economic evaluation^9^. Reality though is that for most, if not all, trials there is no access to IPD and the synthesis has to be based on reported summary findings. Although reported Kaplan-Meier curves for PFS and OS can be digitized and a dataset of “virtual” IPD event-times can be created with the algorithm by Guyot et al., it does not provide the information needed to determine which time-to-progression data point corresponds to which time-to-death data point^10^. Markov-state-transition NMA models have been presented for disease progression^11,12^ and competing risks^13^ based on aggregate level data, but these models assumed constant hazards for transitions between states.

We introduce a method for the joint NMA of PFS and OS that is based on a tri-state (stable, progression, and death) transition model, where time-varying hazard rates and relative treatment effects are modeled with parametric survival functions or fractional polynomials. We illustrate parameter estimation based on aggregate level data.

## 2 Multi-state network meta-analysis framework

### 2.1 Model

At any time *u*, patients in study *i* randomized to treatment arm *k* can be in one of three health states: alive with stable disease (i.e. not progressed), alive and progressed, and dead, with probabilities *S*_*ik*_(*u*), *P*_*ik*_(*u*) and *D*_*ik*_(*u*), respectively, as shown in Figure 1. Let 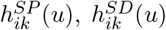, and 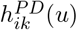 be the hazard rates for the stable-to-progression transition (i.e. disease progression), the stable-to-death transition (i.e. dying pre-progression), and the progression-to-death transition (i.e. dying post-progression).

**Figure 1:**
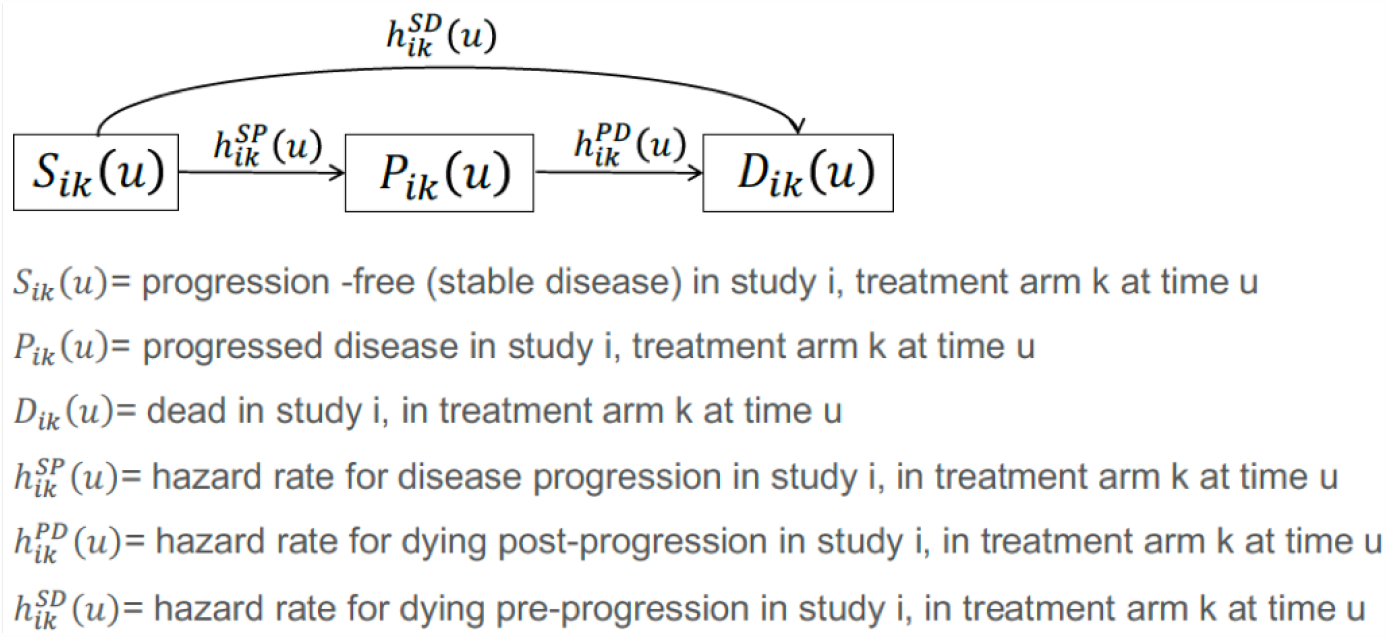
Relationship between stable disease (S), progression (P) and death (D) as used in the multi-state network meta-analysis model.

A multi-state NMA that explicitly estimates each possible transition in a tri-state model and modeling time-varying hazard rates and relative treatment effects with survival functions parameterized as fractional polynomials can be expressed as follows:

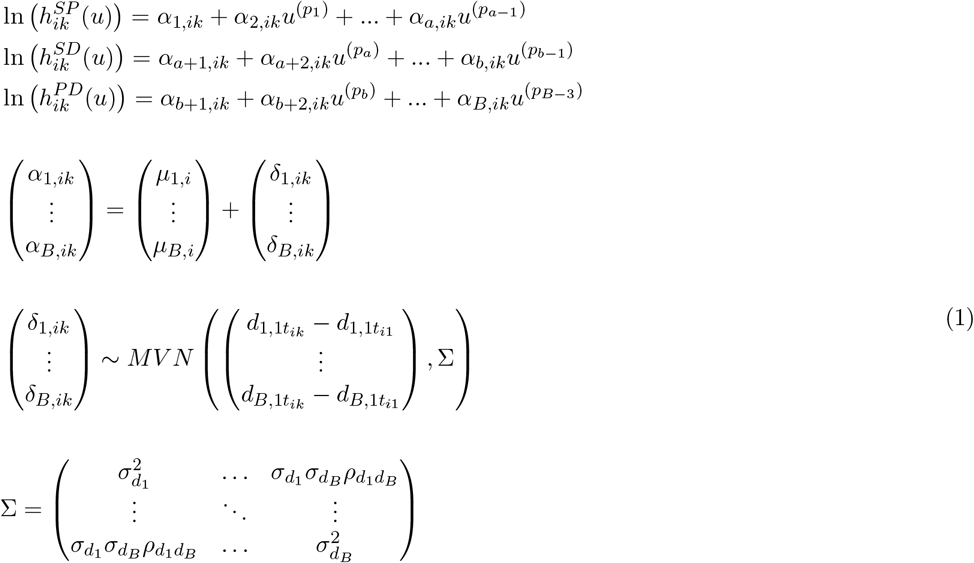

with *δ*_1,*i*1_ = *δ*_1,*i*1_ =, … = *δ*_*B,i*1_ = 0 and *d*_1.11_ = *d*_2.11_ =, …, = *d*_*B*.11_ = 0 for identification.

In Equation 1, *p*_1_, …, *p*_*B*_ are fractional powers and the round bracket notation denotes the Box-Tidwell transformation: *u*^(*p*)^ = *u*^*p*^ if *p* ≠ 0 and *u*^(*p*)^ = ln(*u*) if *p* = 0. Equation 1 also includes the situation of repeated powers, where *p*_*x*_ = *p*_*y*_ for at least 1 pair of indices (*x, y*), 1 ≤ *x < y* ≤ *a* − 1, *a* ≤ *x < y* ≤ *b* − 1, or *b* ≤ *x < y* ≤ *B* − 3. In this situation, 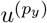 ln(*u*) is used instead of 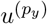 itself. A complete set of flexible fractional polynomials can be created with *p*_1_, …, *p*_*B*−3_ ∈ *{*−2, −1, −0.5, 0, 0.5, 1, 2*}*.

*α*_1,*ik*_, *α*_2,*ik*_, …, *α*_*a,ik*_ are regression coefficients that represent the scale and shape parameters of the log-hazard function describing the stable-to-progression transition in study *i* for study arm *k. α*_*a*+1,*ik*_, *α*_*a*+2,*ik*_, …, *α*_*b,ik*_ are the regression coefficients that represent the log-hazard function for the stable-to-death transition. *α*_*b*+1,*ik*_, *α*_*b*+2,*ik*_, …, *α*_*B,ik*_ are the regression coefficients that represent the scale and shape parameters of the log-hazard function describing the progression-to-death in study *i* for study arm *k*.

When *α*_3,*ik*_ =, …, = *α*_*a,ik*_ = 0, *α*_*a*+3,*ik*_ =, …, = *α*_*b,ik*_ = 0, and *α*_*b*+3,*ik*_ =, …, = *α*_*B,ik*_ = 0 the log-hazard functions for each of the three transitions follow a first order fractional polynomial of which the Weibull and Gompertz are special cases when *p*_1_ = *p*_*a*_ = *p*_*b*_ = 0 and *p*_1_ = *p*_*a*_ = *p*_*b*_ = 1, respectively. When *α*_4,*ik*_ =, …, = *α*_*a,ik*_ = 0, *α*_*a*+4,*ik*_ =, …, = *α*_*b,ik*_ = 0, and *α*_*b*+4,*ik*_ =, …, = *α*_*B,ik*_ = 0 the log-hazard functions for each of the three transitions follow a second-order fractional polynomial.

The *μ*_·,*i*_ reflect the study effects regarding the scale and shape parameters in each study *i*. The *δ*_·,*ik*_ are the study specific true underlying relative treatment effects for the treatment in study arm *k* relative to the treatment in arm 1 of that trial (with *δ*_·,*i*1_ = 0 for identification) regarding the scale and shape parameters of the log-hazard functions for the different transitions, which are modeled with normal distributions with the mean effect for treatment *t* expressed in terms of the overall reference treatment 1, 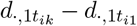, and with a between-study-heterogeneity covariance matrix Σ. 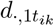 represents the relative treatment effect with treatment *t* in study arm *k* in study *i* relative to reference treatment 1 regarding the scale and shape parameters of the log-hazard functions. We make the assumption of a common between-study correlation 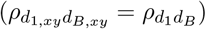 and *σ*_*d*_. represents the common between-study standard deviation.

A fixed effect model is obtained by replacing 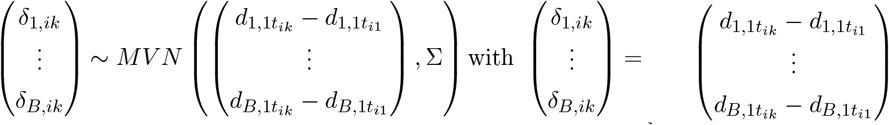.

The random effects model presented with Equation 1 does not account for correlation between trial-specific *δ*_·,*ik*_s in multiple-arm trials (>2 treatments), but can be extended to fit trials with three or more treatment arms by decomposition of a multivariate normal distribution as a series of conditional distributions according to Achana et al.^14^. The conditional distributions for arm *k >* 2, given all arms from 2 to *k* − 1 are:

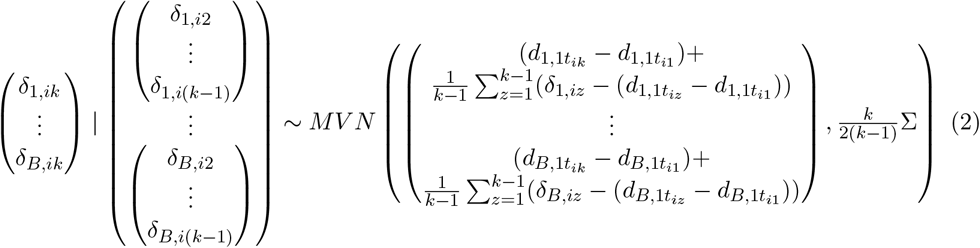

### 2.2 Data and likelihood

For this paper we assume there is no access to IPD for the trials included in the NMA. The parameters will be estimated based on the conditional survival probabilities regarding PFS and OS that can be infered from the published KM curves. (See Appendix A for the algorithm outlining construction of the dataset.) The total follow-up time can be partitioned into *M* successive non-overlapping intervals indexed by *m* = 1, …, *M*. We refer to interval *m* as *U*_*m*_ and write *u* ∈ *U*_*m*_ to denote *u*_*m*_ ≤ *u < u*_*m*+1_. The length of *U*_*m*_ is Δ*u*_*m*_ = *u*_*m*+1_ − *u*_*m*_. For each interval *m*, we propose a binomial likelihood for the conditional survival probabilities regarding PFS and OS at time point *u* relative to the time point at the beginning of the interval *U*_*m*_ according to:

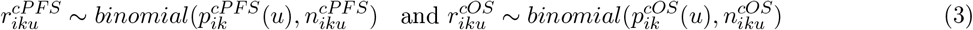

where 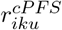 are the observed number of patients who have not yet experienced progression or death at time *u* in the *m*^*th*^ interval in study *i* for treatment arm *k* and 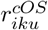 are the observed number of patients who have not died at time *u* in that interval. 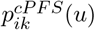 is the underlying conditional survival probability regarding PFS and 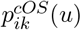 is the underlying conditional survival probability regarding OS, 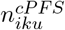 and 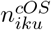 are the corresponding sample sizes at the beginning of the interval. For the *m*^*th*^ interval, the conditional probabilities 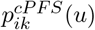 and 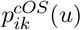 are related to the proportion of patients who are progression free (stable disease) *S*_*ik*_(*u*) and the proportion of patients with progressed disease *P*_*ik*_(*u*) according to:

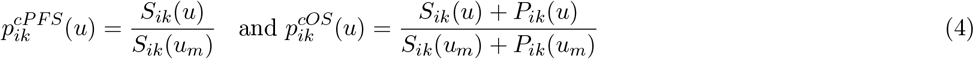

Arbitrary hazard functions can be approximated with a set of discontinuous constant hazard rates over relative short successive time intervals. For each interval *m, S*_*ik*_(*u*), *P*_*ik*_(*u*), and death *D*_*ik*_(*u*) are related to the hazards 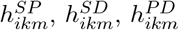 according to the following set of differential equations (See Appendix B):

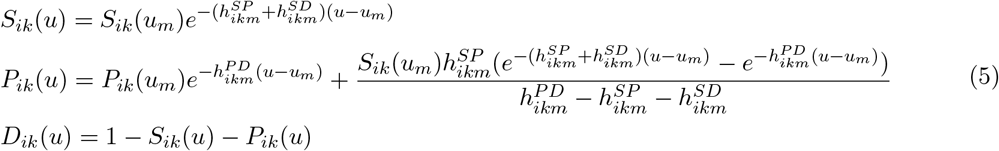

In order to estimate the three parameters 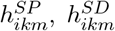, and 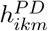 for each interval *m*, we need to define Equation 3, Equation 4 and Equation 5 for at least two time points per interval. In order to improve identifiability of hazard rates in the presence of a small number of events, we use three time points per interval: 1) a time point at 1/3 of the length of the interval 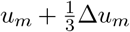, which we define as 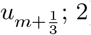) a time point at 2/3 of the length of the interval 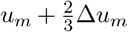, which we define as 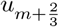; and 3) the time point at the end of the interval *u*_*m*+1_. The obtained estimates of the hazards for interval *m* are assigned to the time point 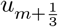 for Equation 1.

## 3 Illustrative example

### 3.1 Evidence base

An example of the multi-state models is presented for a NMA of first line treatment of adult patients with metastatic EGFR+ non-squamous non-small-cell lung cancer (NSCLC) with gefitinib, erlotinib, afatinib, dacomitinib, or platinum-based doublet chemotherapy (PBDC) regimens. Thirteen RCTs were obtained with a systematic literature review (ARCHER1050^15,16^; LUX-LUNG 7^17,18^; LUX-LUNG 3^19,20^; LUX-LUNG 6^20,21^; EURTAC^22–24^; ENSURE^25^; OPTIMAL^26,27^; First-SIGNAL^28^; WJTOG3405^29^; IPASS^30,31^; NEJ002^32,33^; Han2017^34^; Yang2014^35,36^). The evidence network is presented in Figure 2 and the trial-specific PFS and OS curves are provided in the supplementary material (Section C.1).

**Figure 2:**
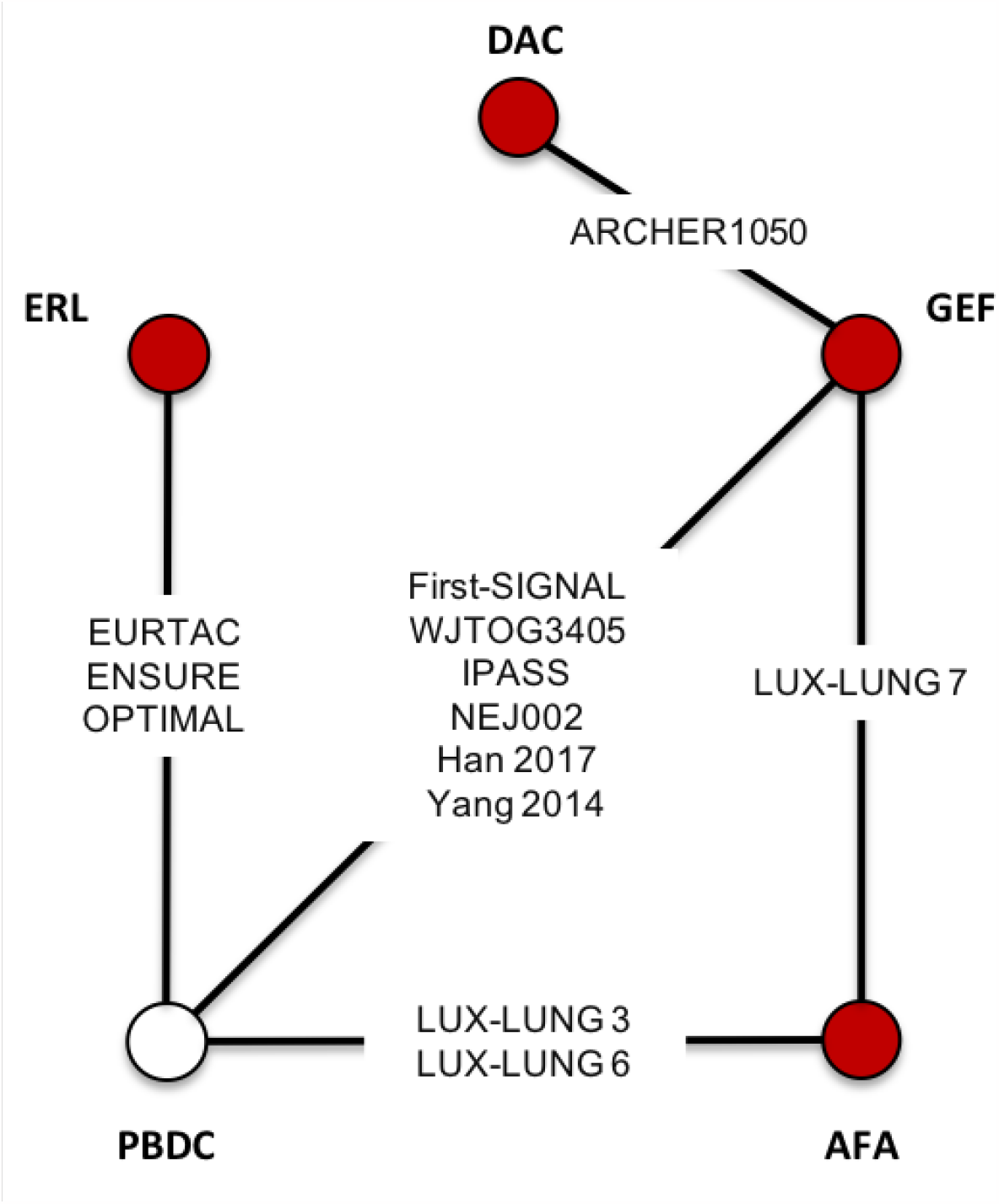
Evidence network of RCTs.

### 3.2 Network meta-analysis to estimate relative treatment effects

The following model was used for the NMAs, which is a simplification of Equation 1 to faciliate parameter estimation, yet believed to be sufficiently flexible to capture the true time-varying hazards for all three transitions in this cancer case study.

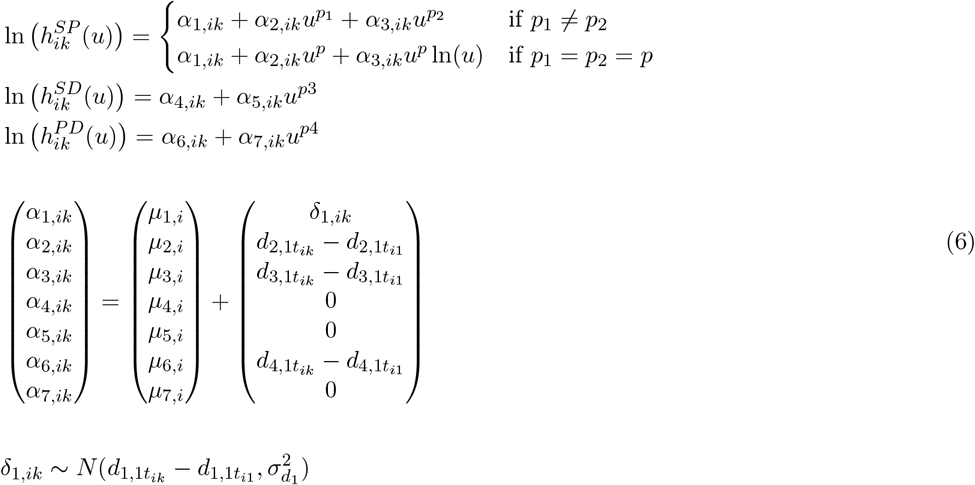

with *u*^0^ = ln(*u*) and *δ*_1,*i*1_ = 0 and *d*_1,11_ = *d*_2,11_ = *d*_3,11_ = *d*_4,11_ = 0 for identification.

When *α*_3,*ik*_ = 0 and *p*_1_ ∈ *{*0, 1*}* the log-hazard functions for the stable-to-progression transition follow a Weibull or Gompertz distribution. When in addition *α*_3,*ik*_ ≠ 0 and *p*_2_ ∈ *{*0, 1*}* the log-hazard functions follow a second order polynomial that are extensions of the Weibull and Gompertz model to allow for arc- and bathtub shaped log-hazard functions. When *α*_5,*ik*_ = 0 the stable-to-death transition follows an exponential distribution. When *α*_5,*ik*_ ≠ 0 and *p*_3_ ∈ *{*0, 1*}* this transition is represented with a Weibull or Gompertz distribution, respectively. When *α*_7,*ik*_ = 0 the log-hazard functions for the progression-to-death transition follow an exponential distribution. When *α*_7,*ik*_ ≠ 0 and *p*_4_ ∈ *{*0, 1*}* this transition is represented with a Weibull or Gompertz distribution.

With this model we assume that the relative treatment effects act on all parameters of the stable-to-progression log-hazard function (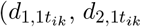, and 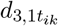). 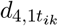 represents the relative treatment effect on the scale parameter of the log-hazard function for the progression-to-death transition. There is one between-study heterogeneity parameter, which is related to the relative treatment effect that acts on the scale of the log-hazard function for the stable-to-progression transition: The *δ*_1,*ik*_ are drawn from a normal distribution with the mean effect for treatment *t* expressed in terms of the overall reference treatment 1, 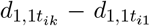, and between study heterogeneity 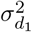. The treatment specific relative effects regarding the first 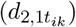 and second 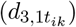 shape parameters of the log-hazard function for the stable-to-progression transition were assumed to be fixed. To accomodate three arm-trials (although not included in this example) Equation 2 can be simplified for this model according to:

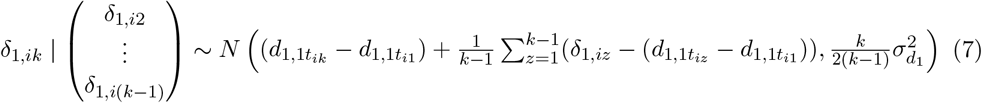

If it is assumed that treatment has only a direct effect on the transitions from stable to progression, the model can be further simplified by setting 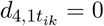.

The following prior distributions for the parameters of the model were used:

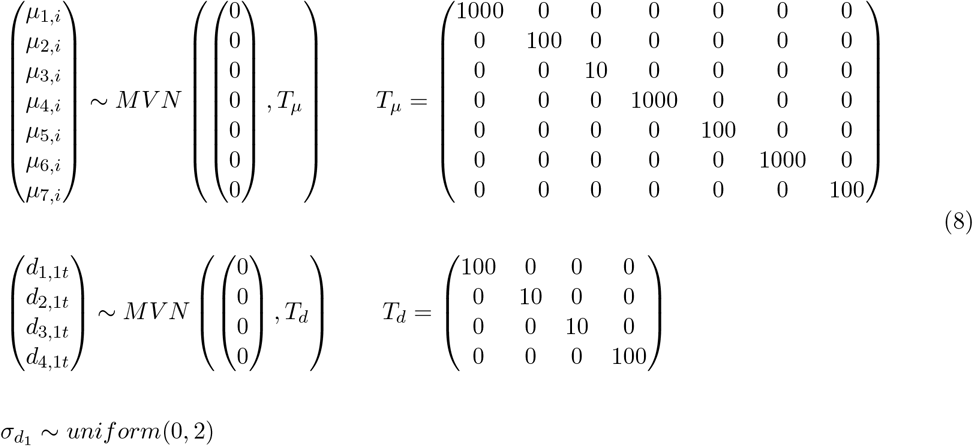

### 3.3 Meta-analysis of absolute effects with overall reference treatment

A NMA provides estimates of relative treatment effects between the competing interventions (i.e. hazard ratios). In order to obtain estimates for the hazard rates over time for the transitions between health states for each treatment, we first need to estimate the time-varying hazard rates for an overall reference treatment, defined as treatment 1, and subsequently apply the hazard ratios of each treatment relative to treatment 1 obtained with the NMA to these baseline hazard rates. As a final step, these time-varying hazard rates for each transition by treatment can be transformed into the distribution *S, P*, and *D* over time, and PFS and OS curves. In this example, gefitinib is defined as treatment 1. Different sources of evidence can be considered for estimating a baseline model, and in the context of cost-effectiveness analysis, it is standard practice to use a (large) observational study that reflects the outcomes in routine practice for the target population of interest. If that is not available, the trial most relevant for the target population can be selected. If multiple studies are deemed relevant then a meta-analysis can be considered. For this example, we still performed a meta-analysis of all gefitinib arms of the trials (instead of selecting one most relevant trial) to illustrate that the proposed framework can also be used to estimate a baseline model if indeed multiple studies are relevant. We used the following fixed effects model:

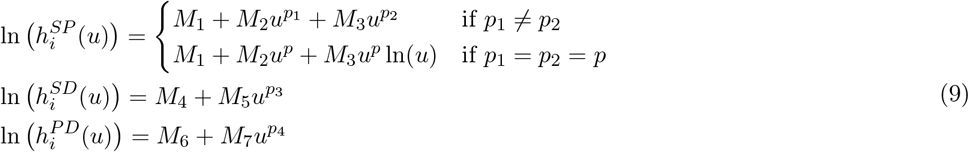

We used the same data structure, likelihood, and link functions as used for the NMA. (See Equation 3, Equation 4, and Equation 5). The prior distribution for this model was:

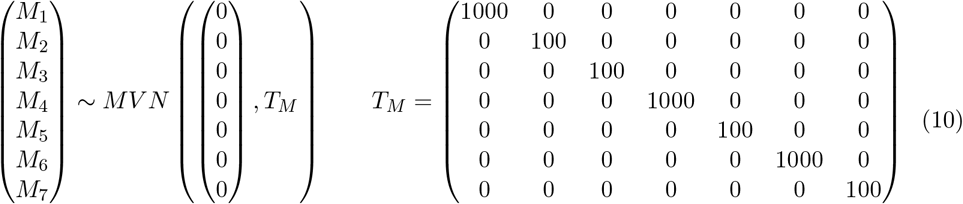

### 3.4 Parameter estimation

The parameters of the different models were estimated using a Markov Chain Monte Carlo (MCMC) method implemented in the JAGS software package^37^. All JAGS analyses were run using the rjags package of R statistical software^38^. See the supplementary material (Section C.2) for the JAGS code for one of the models used to estimate relative treatment effects.

If the sample size or number of PFS or OS related events in interval *U*_*ikm*_ is relatively small, it may be challenging with the MCMC algorithm to distinguish between the “correct answer” for 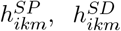, and 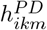, and alternative estimates where either 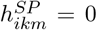 or 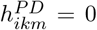. As such, we set a constraint to avoid that 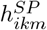 and 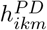 is estimated to be zero. (See the supplementary material Section C.2)

The residual deviance and the deviance information criterion (DIC) were used to compare the goodness-of-fit of the competing models.^39^ The DIC provides a measure of model fit that penalizes model complexity. In general, a more complex model results in a better fit to the data, demonstrating a smaller residual deviance. The model with the better trade-off between fit and parsimony has a lower DIC. A difference of 5 points in the DIC is considered meaningful.^39^

### 3.5 Results

For transparency purposes we first present the results of the meta-analysis of treatment 1 (gefitinib), followed by the results of the NMA, and finally the PFS and OS curves obtained by applying the relative treatment effects obtained with the NMA to the pooled results for treatment 1.

In Table 1, ten competing models for the meta-analysis of treatment 1 that were evaluated are presented. The meta-analysis models with a second order fractional polynomial for the stable-to-progression transition (*SP 2nd order FP(0*.*); SD*..; *PD*.., i.e. *M*_3_ ≠ 0) resulted in the smallest deviance and DICs. The models assuming a Gompertz distribution for this transition (*SP Gompertz; SD*..; *PD*.., i.e. *M*_2_ ≠ 0, 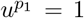, and *M*_3_ = 0) resulted in the largest deviance and DIC. The parameter estimates of a selection of four competing models that show different patterns of time-varying hazards for the three transitions between the health states are presented in Table 2. The actual time-varying hazard rates and corresponding PFS and OS curves with treatment 1 are plotted in Figure 3.

**Table 1:** Model fit criteria for alternative meta-analysis and network meta-analysis models.

| Model | Deviance | pD | DIC |
| --- | --- | --- | --- |
| Meta-analysis Treatment 1 |  |  |  |
| SP 2nd order FP(01); SD Weibull; PD Weibull | 1983 | 7.6 | 1991 |
| SP 2nd order FP(01); SD Weibull; PD exponential | 1983 | 6.7 | 1990 |
| SP 2nd order FP(01); SD exponential; PD Weibull | 2000 | 6.4 | 2007 |
| SP 2nd order FP(01); SD exponential; PD exponential | 2001 | 5 | 2006 |
| SP 2nd order FP(00); SD exponential; PD Weibull | 2018 | 6.8 | 2026 |
| SP Weibull; SD Weibull; PD exponential | 2057 | 5.4 | 2063 |
| SP Weibull; SD exponential; PD Weibull | 2050 | 4.9 | 2056 |
| SP Weibull; SD exponential; PD exponential | 2055 | 5.1 | 2061 |
| SP Gompertz; SD exponential; PD Weibull | 2116 | 4.6 | 2121 |
| SP Gompertz; SD exponential; PD exponential | 2155 | 4.8 | 2161 |
| Network meta-analysis |  |  |  |
| SP 2nd order FP(01) FE3; SD Weibull; PD Weibull FE1(scale) | 5849 | 119 | 5968 |
| SP 2nd order FP(01) FE3; SD Weibull; PD exponential FE | 5864 | 108 | 5972 |
| SP 2nd order FP(01) FE3; SD Weibull; PD exponential | 5872 | 98 | 5970 |
| SP 2nd order FP(01) FE3; SD exponential; PD Weibull FE1(scale) | 5943 | 91 | 6034 |
| SP 2nd order FP(01) FE3; SD exponential; PD Weibull | 5970 | 91 | 6061 |
| SP 2nd order FP(01) FE3; SD exponential; PD exponential | 5980 | 79 | 6059 |
| SP Weibull FE2; SD Weibull; PD exponential FE | 5977 | 75 | 6052 |
| SP Weibull FE2; SD exponential; PD Weibull FE1(scale) | 6005 | 81 | 6087 |
| SP Weibull FE2; SD exponential; PD Weibull | 6055 | 71 | 6126 |
| SP Weibull FE2; SD exponential; PD exponential | 6069 | 61 | 6130 |
| SP 2nd order FP(01) RE3; SD Weibull; PD Weibull FE1(scale) | 5803 | 129 | 5932 |
| SP 2nd order FP(01) RE3; SD Weibull; PD exponential FE | 5821 | 116 | 5937 |
| SP 2nd order FP(01) RE3; SD exponential; PD Weibull FE1(scale) | 5898 | 111 | 6009 |
| SP Weibull RE2; SD exponential; PD Weibull FE1(scale) | 5977 | 86 | 6063 |

Deviance: defined as $-2 \times \log(\text{likelihood})$
DIC: Deviance information criterion, $DIC = \bar{D} + pD$ . $\bar{D}$ is the posterior mean of the deviance
pD: the posterior mean of the deviance minus the deviance of the posterior means, corresponds to the effective number of parameters
SP: stable-to-progression transition; SD: stable-to-death transition; PD: progression-to-death transition; FP(01) 2nd order fractional polynomial with $p_1 = 0$ and $p_2 = 1$ ; FP(00) 2nd order fractional polynomial with $p_1 = p_2 = 0$ ; FE3 and RE3: fixed and random effects model with relative treatment effects impacting the three parameters of the 2nd order fractional polynomial log-hazard function for the stable-to-progression transition; FE2 and RE2: fixed and random effects model with relative treatment effects impacting the scale and shape parameters of the Weibull or Gompertz log-hazard function for the stable-to-progression transition; FE1(scale): fixed effects model with relative treatment effect impacting the scale parameters of the Weibull or Gompertz log-hazard function for the progression-to-death transition; FE: fixed effects model with relative treatment effect impacting the rate parameter of the exponential log-hazard function for the progression-to-death transition.

**Table 2:** Parameter estimates regarding hazard rates over time for the stable-to-progression transition, the stable-to-death transition, and progression-to-death transition with treatment 1 for a selection of alternative meta-analysis models.

| Parameter | Model 1 |  |  | Model 2 |  |  | Model 3 |  |  | Model 4 |  |  |
| --- | --- | --- | --- | --- | --- | --- | --- | --- | --- | --- | --- | --- |
|  | estimate | low | high | estimate | low | high | estimate | low | high | estimate | low | high |
| $M_1$ | -3.752 | -4.081 | -3.512 | -3.758 | -4.102 | -3.514 | -3.628 | -3.777 | -3.469 | -3.136 | -3.23 | -3.041 |
| $M_2$ | 0.974 | 0.768 | 1.241 | 0.963 | 0.761 | 1.237 | 0.512 | 0.441 | 0.583 | 0.052 | 0.044 | 0.06 |
| $M_3$ | -0.073 | -0.101 | -0.049 | -0.069 | -0.098 | -0.046 | | | | | | |
| $M_4$ | -4.786 | -5.306 | -4.378 | -4.808 | -5.34 | -4.385 | -4.587 | -4.605 | -4.513 | -4.589 | -4.605 | -4.527 |
| $M_5$ | -0.984 | -1.17 | -0.578 | -0.992 | -1.17 | -0.615 | 0.013 | -0.013 | 0.07 | | | |
| $M_6$ | -2.983 | -3.431 | -2.504 | -2.779 | -2.872 | -2.681 | -2.968 | -3.059 | -2.879 | -2.97 | -3.053 | -2.881 |
| $M_7$ | 0.071 | -0.092 | 0.227 | | | | | | | | | |

Model 1: SP 2nd order FP(01); SD Weibull; PD Weibull
Model 2: SP 2nd order FP(01); SD Weibull; PD exponential
Model 3: SP Weibull; SD Weibull; PD exponential
Model 4: SP Gompertz; SD exponential; PD exponential
estimate: the median of the posterior distribution; low and high: lower and upper bound of the 95% credible interval corresponding to the 2.5th and 97.5th percentile of the posterior distribution.

**Figure 3:**
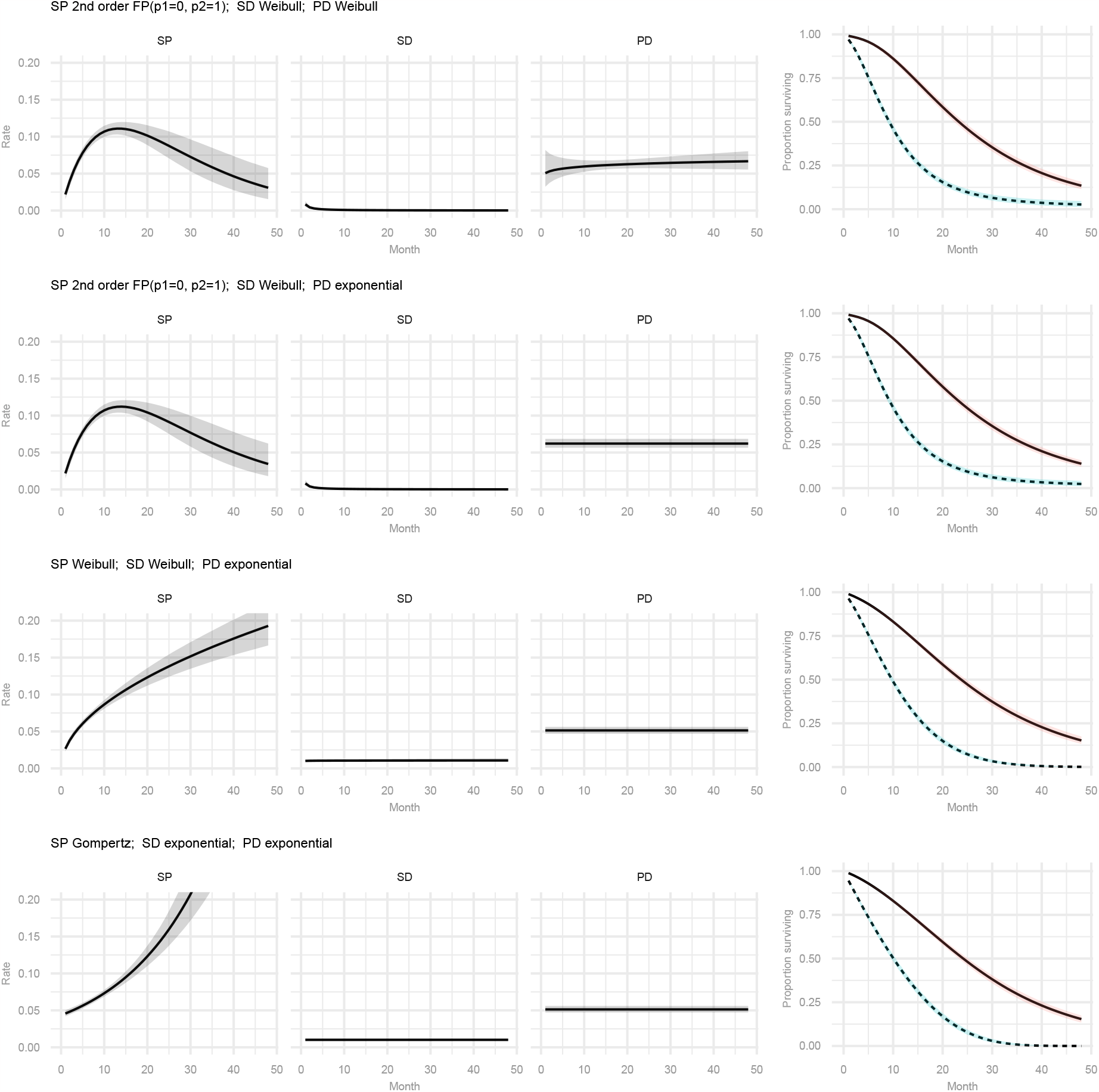
**Pooled estimates of hazard rates over time for the stable-to-progression transition (SP), stable-to-death transition (SD), and progression-to-death transition (PD), and PFS and OS curves with treatment 1 from a selection of alternative multi-state fixed effects meta-analysis models**

For the NMA, 14 competing models were evaluated; ten fixed effects models and four random effects models. (See Table 1) The NMA models that assumed a second order fractional polynomial for the stable-to-progression transition had a lower deviance and DIC than the corresponding simpler models assuming a Weibull distribution for this transition. The models with a Weibull distribution for the stable-to-death transition had a lower DIC than the corresponding models assuming an exponential distribution for this transition. Models with a Weibull distribution for the progression-to-death transition were not a meaningful improvement over the corresponding models that assumed an exponential distribution for this transition. Comparing models that assumed a relative treatment effect for the progression-to-death transition with the corresponding models without this relative treatment effect indicates that a relative effect may be a relevant component to include for this transition in some models. The second order fractional polynomial and Weibull random effects models performed better than their fixed effects equivalents, indicating that incorporating between-study heterogeneity is important.

Parameter estimates for the random effects second order fractional polynomial model (*SP 2nd order FP(01) RE3; SD Weibull; PD Weibull FE1(scale)*), the random effects Weibull model (*SP Weibull RE2; SD exponential; PD Weibull FE1(scale)*, and two fixed effects models (*SP Weibull FE2; SD Weibull; PD exponential FE* and *SP Weibull FE2; SD exponential; PD exponential*) are presented in Table 3. The corresponding time-varying HRs with each treatment relative to treatment 1 for the stable to progression transitions are presented in Figure 4, and the constant HRs for the progression-to-death transition in Figure 5. (Please note that we did not assume a relative treatment effect for the stable-to-death transition). Applying the relative treatment effect parameter estimates describing the HRs over time obtained with these NMA models to the parameter estimates of the models used for the analysis of treatment 1, we obtain the PFS and OS curves by treatment, as presented in Figure 6. In order to illustrate the width of the 95% credible intervals of these survival curves due to the uncertainty in the time-varying HRs, we ignored the uncertainty for the reference treatment 1. (If the uncertainty of the meta-analysis would have been incorporated as well, the 95% credible intervals would have been a bit wider.)

**Table 3:** Relative treatment effect parameters regarding time-varying hazard rates for the stable-to-progression transition, stable-to-death transition, and progression-to-death transition for a selection of alternative network meta-analysis models.

| Parameter | Model 1 |  |  | Model 2 |  |  | Model 3 |  |  | Model 4 |  |  |
| --- | --- | --- | --- | --- | --- | --- | --- | --- | --- | --- | --- | --- |
|  | estimate | low | high | estimate | low | high | estimate | low | high | estimate | low | high |
| $d_{1,11}$ | 0 | 0 | 0 | 0 | 0 | 0 | 0 | 0 | 0 | 0 | 0 | 0 |
| $d_{1,12}$ | -0.661 | -1.686 | 0.286 | -0.483 | -1.299 | 0.299 | -0.481 | -1.013 | 0 | -0.531 | -1.1 | -0.048 |
| $d_{1,13}$ | -0.096 | -0.904 | 0.709 | -0.131 | -0.74 | 0.485 | -0.069 | -0.36 | 0.237 | -0.14 | -0.433 | 0.17 |
| $d_{1,14}$ | 0.222 | -1.019 | 1.389 | -0.019 | -0.981 | 0.977 | 0.016 | -0.41 | 0.445 | 0.042 | -0.388 | 0.457 |
| $d_{1,15}$ | 0.7 | 0.137 | 1.254 | 0.636 | 0.208 | 1.056 | 0.685 | 0.449 | 0.932 | 0.689 | 0.459 | 0.918 |
| $d_{2,11}$ | 0 | 0 | 0 | 0 | 0 | 0 | 0 | 0 | 0 | 0 | 0 | 0 |
| $d_{2,12}$ | -0.84 | -1.635 | -0.114 | -0.152 | -0.411 | 0.127 | -0.085 | -0.349 | 0.187 | -0.019 | -0.283 | 0.262 |
| $d_{2,13}$ | -0.206 | -0.645 | 0.169 | -0.059 | -0.209 | 0.089 | -0.074 | -0.227 | 0.074 | -0.045 | -0.199 | 0.101 |
| $d_{2,14}$ | -0.547 | -1.089 | -0.013 | -0.341 | -0.535 | -0.135 | -0.347 | -0.539 | -0.156 | -0.344 | -0.536 | -0.15 |
| $d_{2,15}$ | 0.127 | -0.256 | 0.492 | 0.122 | -0.005 | 0.249 | 0.156 | 0.026 | 0.28 | 0.084 | -0.035 | 0.209 |
| $d_{3,11}$ | 0 | 0 | 0 | | | | | | | | | |
| $d_{3,12}$ | 0.217 | 0.053 | 0.388 | | | | | | | | | |
| $d_{3,13}$ | 0.024 | -0.028 | 0.079 | | | | | | | | | |
| $d_{3,14}$ | 0.029 | -0.042 | 0.096 | | | | | | | | | |
| $d_{3,15}$ | -0.021 | -0.093 | 0.049 | | | | | | | | | |
| $d_{4,11}$ | 0 | 0 | 0 | 0 | 0 | 0 | 0 | 0 | 0 | | | |
| $d_{4,12}$ | -0.114 | -0.997 | 0.37 | 0.339 | 0.003 | 0.696 | 0.046 | -0.462 | 0.503 | | | |
| $d_{4,13}$ | -0.041 | -0.273 | 0.181 | 0.051 | -0.135 | 0.247 | -0.134 | -0.333 | 0.074 | | | |
| $d_{4,14}$ | 0.24 | -0.094 | 0.564 | 0.146 | -0.18 | 0.48 | 0.207 | -0.105 | 0.516 | | | |
| $d_{4,15}$ | -0.245 | -0.393 | -0.095 | -0.299 | -0.45 | -0.148 | -0.273 | -0.43 | -0.112 | | | |
| $\sigma_{d_1}$ | 0.445 | 0.237 | 0.891 | 0.372 | 0.211 | 0.749 | | | | | | |

Model 1: SP 2nd order FP(01) RE3; SD Weibull; PD Weibull FE1(scale)
Model 2: SP Weibull RE2; SD exponential; PD Weibull FE1(scale)
Model 3: SP Weibull FE2; SD Weibull; PD exponential FE
Model 4: SP Weibull FE2; SD exponential; PD exponential
estimate: the median of the posterior distribution. low and high: lower and upper bound of the 95% credible interval corresponding to the 2.5th and 97.5th percentile of the posterior distribution. $d_{1,1t}$ : relative treatment effect with treatment $t$ versus treatment 1 regarding the scale of the log-hazard function describing the stable-to-progression transition. $d_{2,1t}$ : relative treatment effect with treatment $t$ versus treatment 1 regarding the first shape parameter of the log-hazard function describing the stable-to-progression transition. $d_{3,1t}$ : relative treatment effect with treatment $t$ versus treatment 1 regarding the second shape parameter of the log-hazard function describing the stable-to-progression transition. $d_{4,1t}$ : relative treatment effect with treatment $t$ versus treatment 1 regarding the scale of the log-hazard function describing the progression-to-death transition

**Figure 4:**
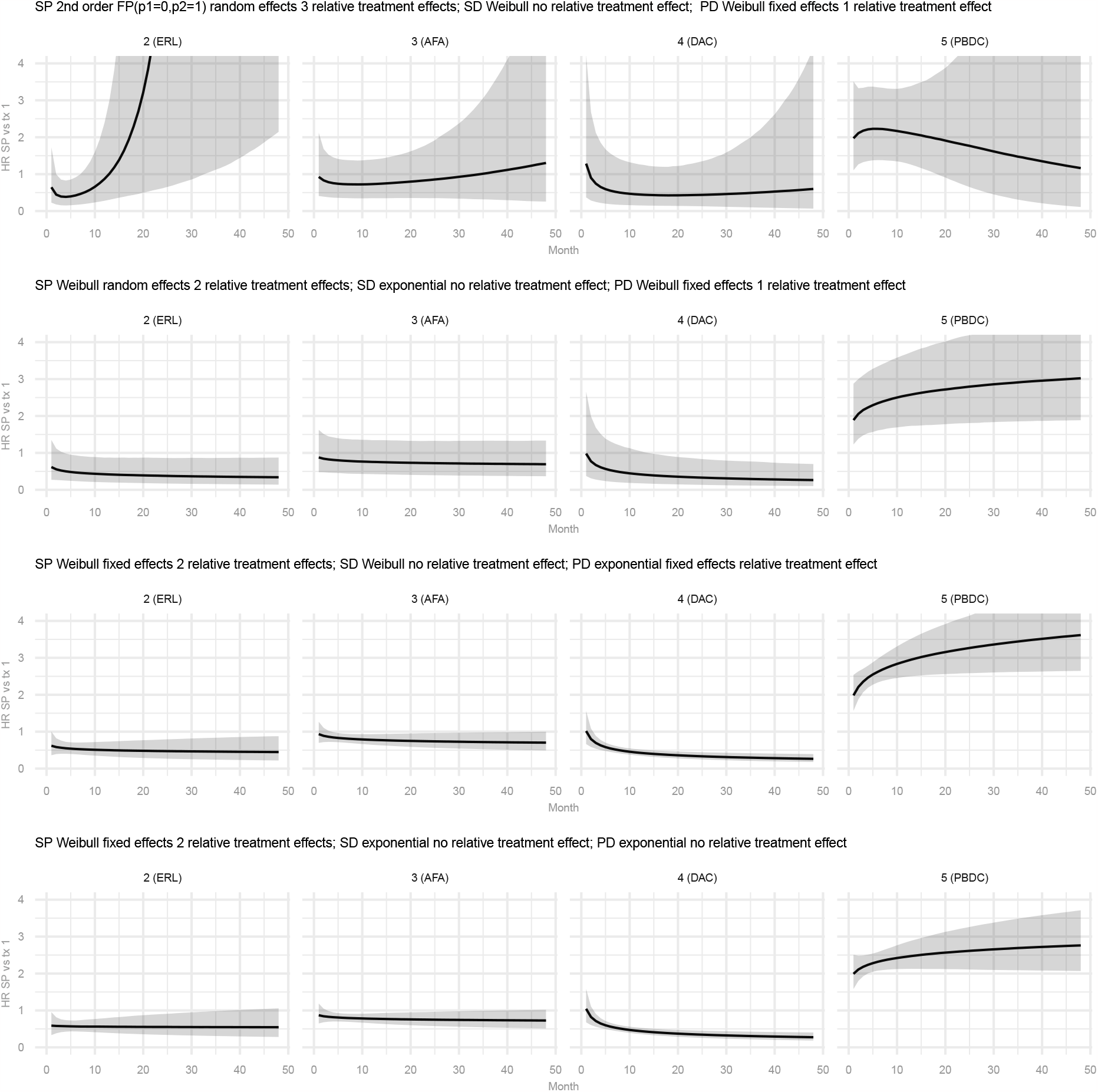
Estimates of hazard ratios for the stable-to-progression transition with treatments 2-5 relative to treatment 1 from a selection of alternative multi-state network meta-analysis models.

**Figure 5:**
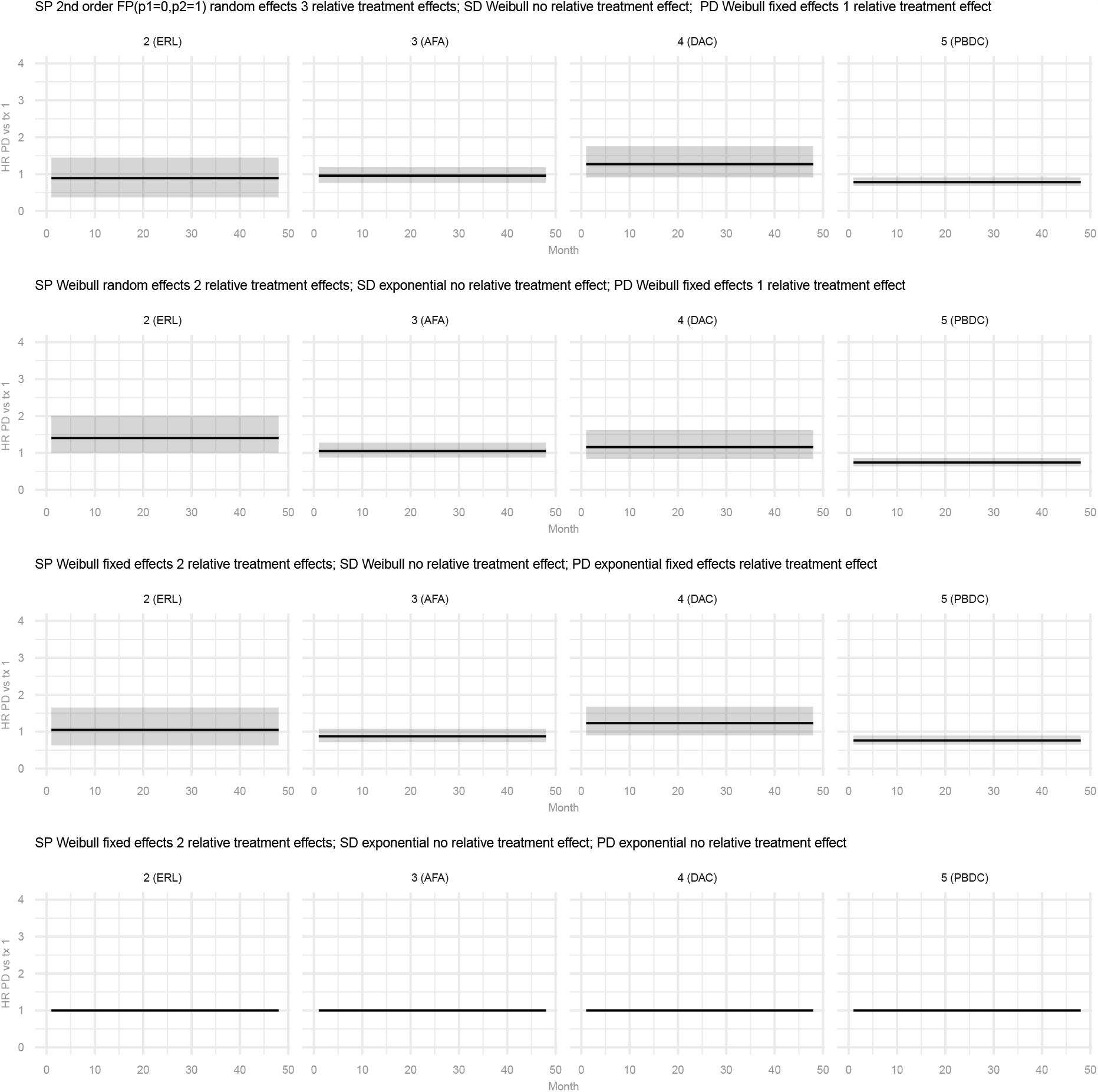
Estimates of hazard ratios for the progression-to-death transition with treatments 2-5 relative to treatment 1 from a selection of alternative multi-state network meta-analysis models.

**Figure 6:**
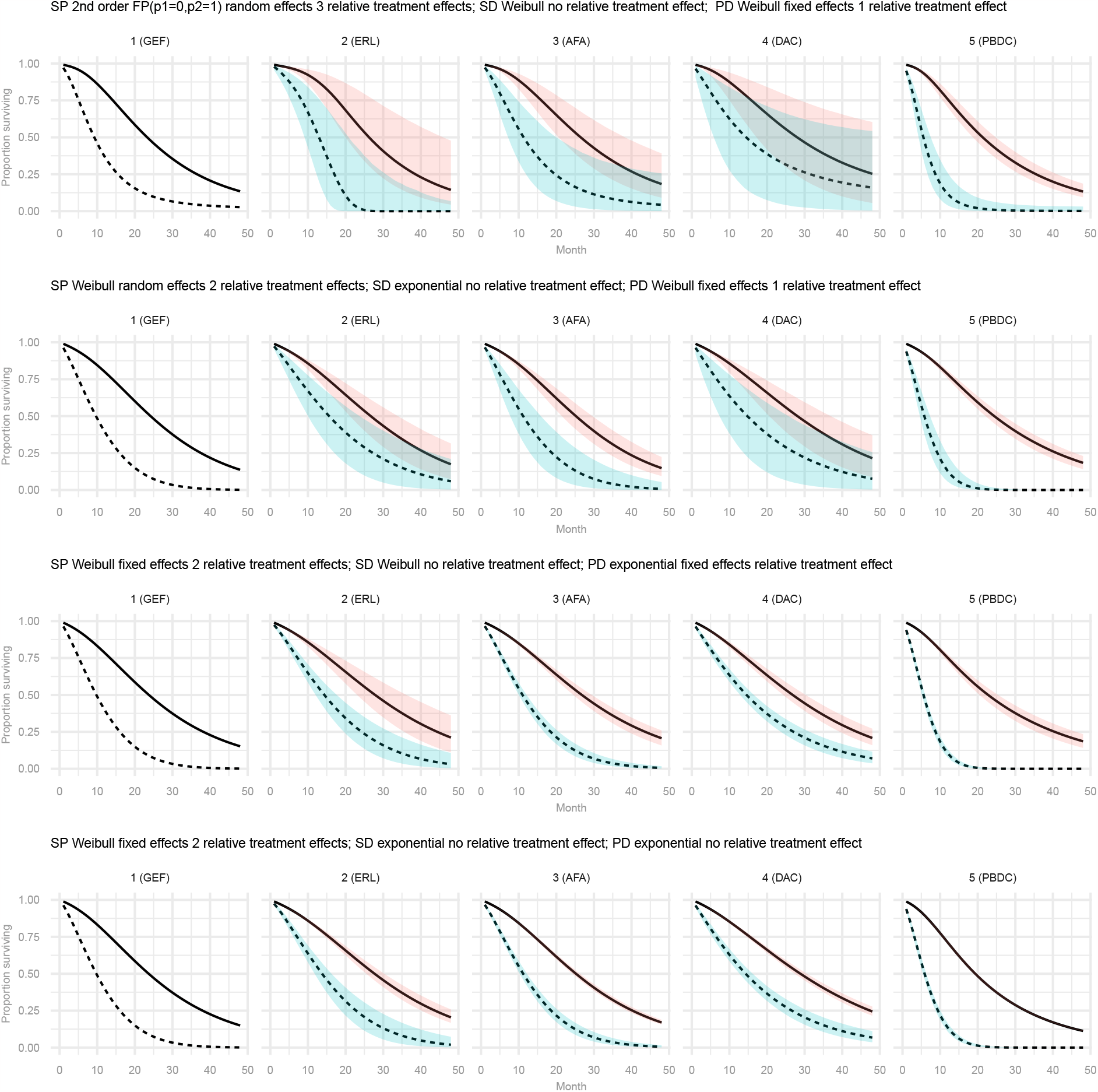
Estimates of progression-free survival and overall survival for treatment 1-5 obtained with a selection of alternative multi-state network meta-analysis models.

## 4 Discussion

With this paper we present a method for the joint NMA of PFS and OS that is based on a tri-state transition model. This method extends existing parametric NMA methods for time-to-event data^4–7^ by defining the structural relationship between PFS and OS according to the stable, progression, and death states that define the course of disease over time. Instead of modeling the time-varying hazard rates for PFS and OS separately, we model the time-varying transition rates between the three health states simultaneously. The primary advantage of this evidence synthesis framework is that estimates for PFS and OS remain consistent over time, which is needed for decision and economic modeling.

The primary reason to propose the method described in this paper is to facilitate parameterization of multi-state cost-effectiveness models based on summary level data. In order to do so, we describe the conditional survival probability for PFS and OS with two separate binomial likelihoods for a given interval *m* (See Equation 3), and capture their relationship with Equation 4 and Equation 5. The implication of this approach is that we assume that 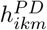 is independent from 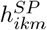, or 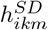, or both 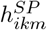 and 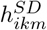. The correlation between conditional PFS and conditional OS is explained by shared parameters 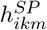 and 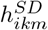. For the purposes of decision making, the uncertainty in estimates is at least as important as the point estimates themselves. If the structural assumption regarding the relationship between the likelihoods for conditional PFS and OS does not capture their correlation appropriately, the precision in some or all of the transition rates, and therefore OS, will be overestimated. We performed a simple simulation to assess the performance of estimating the hazard rates for the three transitions in a given time interval using the three conditional PFS and OS data points for that interval. Overall, the coverage probabilities of the 95% credible intervals for the hazard rates are acceptable. (See Section C.4 for more detail.) However, more elaborate simulation studies are recommended to investigate this in more detail.

To estimate the model parameters, we opted to use three conditional PFS and three conditional OS data points for each time interval. In principle, two conditional PFS and OS data points (i.e. four data points in total) would be sufficient to estimate the three constant hazard rates corresponding to the possible transitions in a given interval. When the number of events are small, however, the MCMC algortithm may not be able to distinguish between the “correct solutions” for the three hazard rates and alternative solutions corresponding to a situation where either 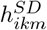 or 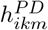 equates zero. By using three conditional PFS and OS data points for each time interval, this is less likely to be the case. (In our example, we also used a likelihood constraint to avoid 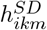 or 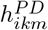 equate zero.) Using three conditional PFS and OS data points for each time interval implied 3-month constant hazards in our example, longer than if two conditional PFS and OS data points would have been used (i.e. 2-month constant hazards). In principle, we want intervals sufficiently short to allow the models to reflect true changes in the hazards over time. However, the shorter the interval, the smaller the number of transitions in an interval and the greater the uncertainty of the interval-specific hazard rates. With our approach, the subsequent interval-specific rates are “connected” with the model thereby improving interval-specific estimates, but the uncertainty in scale and shape parameter estimates will be smaller when interval-specific estimates can be estimated with greater precision. In essence with longer time intervals we remove some of the parameter uncertainty in exchange for a greater structural assumption. As a first attempt in assessing the impact of the length of the interval on model estimates, we performed a comparison of the estimated time-varying hazards for the different transitions as obtained with the meta-analysis of treatment 1 according to Model 1 based on an interval length of 3 months using three data points (as used in the example analyses) and an interval length of 1.5 months also using three data points. (See supplementary material, Section C.5) Out of the presented models for the example, Model 1 is the most flexible in terms of capturing changes in the hazards over time and therefore best suited to compare the impact of the length of the time-interval. As depicted, the estimated hazards and patterns are similar between the two analyses, indicating that the method seems robust to the chosen interval length, at least for intervals between 1.5 and 3 months. That being said, further research is needed to define the optimal length of intervals and number of data points to use per interval given the observed number of events in an interval, the population size at risk at the beginning of the interval, and rate of change of the hazards over time.

Given the flexibility of the proposed framework, decisions need to be made regarding potentially relevant model structures for its implementation for a specific study. For the illustrative example, we made model choices that we considered appropriate for a typical cancer case study where treatment aims to prolong survival. First, we assumed that a relative treatment effect for the stable-to-death transition was not needed reflecting the belief that differences in survival between treatments are only due to differences in delayed or avoided tumor progression, and not due to other treatment-related (adverse) events. Second, we assumed that a function more complex than a first-order fractional polynomial with a constant relative treatment effect was not required for the progression-to-death transition because this transition is conditional upon experiencing progression and modeled in relation to follow-up time. One could even argue that a relative treatment effect for this transition is not needed when treatment is discontinued upon progression. However, the DIC indicated that adding this parameter to the models resulted in a meaningful improvement for the example analyses, which is primarily related to the PBDC trials. Reasons to include a treatment effect parameter for the progression-to-death transition is treatment cross-over upon progression in a subset of trials, if post-progression treatment between trial arms differ, or if the post-progression mortality patterns are expected to differ between treatments due to other reasons. Furthermore, there may be correlations between the time-to-progression and subsequent time-to-death that could induce treatment effects for the progression-to-death transition. For these reasons exploring incorporating a treatment effect for the progression-to-death transition should be explored. Third, out of the possible fractional polynomials, we only evaluated exponential, Weibull and Gompertz models and their extensions where the additional parameter related the log-hazard to time or log-time. We did not consider any of the negative power transformations of time, primarily because these functions do not link to known survival distributions and the second-order models we did use have already the flexibility to capture arc-shaped hazard functions.

This brings us to the point of model selection in general. Factors to consider when defining a relevant subset of competing models available within the proposed framework include: (i) the required flexibility to capture time-related patterns of the hazard functions for the different transitions; (ii) which transitions do we expect to vary by treatment; (iii) does treatment only impact scale or also shape parameters; (iv) how do we incorporate between-study heterogeneity; and (v) availability of data in relation to the number of parameters to estimate. The degree of flexibility of the competing log-hazards functions is arguably most important for the stable-to-progression transition as these hazards may vary substantially over time and between treatments. For the progression-to-death transition, the stability of estimates is arguably the most important given the potential need for overall survival extrapolation. For the stable-to-death transition, the importance of the appropriate function depends on the expected hazard rates in comparison to the rates for the other transitions. If the stable-to-death transition rates are relatively low, model misspecification may have a limited impact. In addition, as mentioned, these transitions may reflect background mortality and, as such, we do not need treatment effect-parameters. When selecting the preferred models out of the defined set of competing models of potential relevance, we can use DIC to inform model selection, as we did in our example, but estimating the total residual deviance and comparing this to the number of data points is a useful addition as it tells us how well the models are fitting the data. In general, evaluating all possible competing models of relevance regarding fit to the data may not be feasible from a practical perspective; even if we are estimating DIC using only a small number of exploratory MCMC samples, the computational burden is still substantial. Future research is recommended to inform a model selection strategy or algorithm that results in a set of models that is likely to cover the distribution of transition rates between the health states, results in realistic extrapolations over time, and, given the computational burden of the more complex models for large datasets, can be evaluated in a reasonable amount of time.

The proposed evidence synthesis framework relates directly to clock forward time-inhomogeneous Markov models where treatment specific transition rates between health states are only a function of time in the model. A frequently used approach for cost-effectiveness analysis of cancer treatments are partitioned survival models. However, the main limitation is that extrapolated parametric PFS and OS curves for a given treatment may cross. This will not be case with Markov models and, as such, are preferred as long as time-varying transition rates between health states can be estimated that reflect the actual PFS and OS of the treatments compared. As far as we know, the method presented in this paper is the first to facilitate this based on reported aggregate level data. In order to obtain the parameter estimates for a cost-effectiveness analysis we need to define a baseline model and a NMA model. The baseline model provides estimates for the absolute effect with the reference treatment, which in this case are the time-varying log-hazard rates between each of the three health states. The NMA model provides estimates of the relative treatment effects of each intervention in the network relative to the reference treatment, which in this case are the time-varying log hazards ratios. The absolute effect with each treatment is obtained by adding the relative treatment effects from the NMA to the absolute effect with the reference treatment from the baseline model, and subsequently transforming these to the natural scale by inverting the log-link function^40^. In the current example we used the RCT evidence base to estimate the baseline meta-analysis model as well as the NMA model. However, for an actual cost-effectiveness analysis it is recommended using an evidence base that reflects expected outcomes with the reference treatment for the target population in routine practice for the baseline model, preferably a large long-term routine practice observational study. If that is not available, a meta-analysis of the most relevant or recent trials can be considered.

The estimates obtained with the proposed evidence synthesis models can also be used in semi-Markov individual simulation models (i.e. models where some transitions are affected by time in an intermediate state). For example, imagine a cost-effectiveness model of first-line cancer treatment consisting of the three health states stable, progression, and death. The stable-to-progression and stable-to-death transitions can be estimated based on first-line trials using the proposed multi-state (network) meta-analysis method. The progression-to-death transitions in these trials no longer represent current standard of care and we need to estimate these transition rates based on overall survival data from second line trials of current treatment using a separate (network) meta-analysis model. Similarly, we can use the proposed evidence synthesis models in the context of semi-Markov simulation models describing sequential cancer treatment. Imagine a model consisting of four health states: 1) stable disease with first line treatment, 2) progression with first line treatment/stable disease with second line treatment, 3) progression with second line treatment, and 4) death. First-line treatment transitions from stable-to-progression and stable-to-death are estimated with one multi-state (network) meta-analysis model based on first line trials, and the second-line treatment transitions from stable-to-progression, stable-to-death, and progression to death (reflecting third line treatment and beyond) are estimated with another multi-state (network) meta-analysis model based on second line trials. In general, for the transitions in a simulation model for which the “clock is reset” a separate multi-state (network) meta-analysis needs to be performed.

Time-varying transition rates from an intermediate health state, e.g. progressed disease, are typically modeled as a function of time since entering that state using (time-reset) semi Markov models. For clock-forward Markov models, where transition rates are only a function of time in the model, typically constant transition rates are used for the transitions from intermediate health states. However, we want to highlight that this is not a requirement and time-varying transition rates in a clock-forward Markov model can be defended in certain situations. For example, a monotonically decreasing hazard function (corresponding to Weibull distribution) for the progression-to-death transition means that an individual progressing after (say) 6 months has a greater probability of dying in the subsequent month than (say) an individual who progressed after 24 months. This reflects the possible scenario that more severe individuals or individuals without any treatment response are more likely to die faster once progressed than less severe patients who did show an initial response and progressed slower. In fact, this is a potential benefit of this multi-state evidence synthesis method. Separate estimation of transition rates between health states cannot capture this aspect. To capture differential patterns between treatments, a relative treatment effect for the progression-to-death transition can be incorporated in the evidence synthesis model.

All studies provided PFS and OS Kaplan-Meier data in the example analyses. In principle, the NMA model can be extended to create a shared-parameter model to incorporate studies that only provide information for PFS or only for OS. Studies with only PFS data provide evidence regarding the stable-to-progression and stable-to-death transitions and contribute to estimating the corresponding treatment specific hazard ratios if these are assumed fixed or exchangeable across all studies providing direct or indirect evidence for that particular intervention. (When a meta-analysis of absolute effects with the overall reference treatment is performed, the fixed effects or exchangeability assumption applies to the transition rates.) Incorporating studies that only provide OS data for a particular intervention in the NMA will require the additional assumption of fixed or exchangeable rates for one of the transitions across all studies for that intervention, if treatment is assumed to impact more than just the stable-to-progression transition in order to facilitate parameter estimation. A related topic for future research is whether and how this framework can be used to validate PFS as a surrogate for OS and to predict OS for novel interventions for which only mature PFS is available. This will be of great benefit for cost-effectiveness analyses.

## 5 Conclusion

We introduced a method for the joint meta-analysis of PFS and OS that is based on a non-homogenous Markovian tri-state transition model. Arbitrary hazard rate functions can be approximated by piecewise constant hazard rates at successive time intervals, and are flexibly modeled as (fractional) polynomial functions of time. The proposed approach relaxes the proportional hazards assumption, extends to a network of more than two treatments, and simplifies the parameterization of decision and cost-effectiveness analyses. The data needed to run these analyses can be extracted directly from published survival curves.

## Data Availability

Dataset is available upon reasonable request.

## 6 Conflict of interest

The authors have no conflict of interest related to this work to report.

## 7 Funding

JP Jansen was supported by the UCSF Academic Senate Committee on Research (Academic Senate RAP Grant). TA Trikalinos was supported in part by a grant from the National Cancer Institute (5U01CA265750). The funders had no role in the preparation, review, or approval of the manuscript or the decision to submit the manuscript for publication.

## 8 Data availability statement

The data that support the findings of this study are available from the corresponding author upon reasonable request.

## Appendices

### A Constructing dataset for analyses

Data inputs required are the coordinates extracted from digitally scanned PFS and OS Kaplan-Meier curves: time points (*u*),corresponding survival probabilities (*s*_*u*_), and corresponding population size at risk (*n*_*u*_). These points must capture all steps in the curve, and may require adjustments to the extracted coordinates to ensure the survival probabilities are decreasing with time. For both curves it should include the times at which numbers at risk are reported below the curve.

The total follow-up time can be partitioned into *M* successive non-overlapping intervals indexed by *m* = 1, …, *M*. We refer to interval *m* as *U*_*m*_ and write *u* ∈ *U*_*m*_ to denote *u*_*m*_ ≤ *u < u*_*m*+1_. The length of *U*_*m*_ is Δ*u*_*m*_ = *u*_*m*+1_ − *u*_*m*_. For each time interval *m*, we want to obtain four data points: At the beginning of the interval, *u*_*m*_; at 1/3 of the length of the interval, 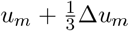, which we define as 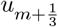; at 2/3 of the length of the interval, 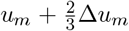, which we define as 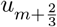; and at the end of the interval, *u*_*m*+1_. It is desirable to have the time intervals defined in such a way that (some) of these time points are aligned with the time point for which the size of the at-risk population is reported below the published Kaplan-Meier curves, and are the same for PFS and OS where available. For the current study, we used intervals with a length of three months.

If no PFS or OS proportion have been recorded for a specific time point of interest (i.e. whole months), a corresponding value for *s*_*u*_ can be obtained by linear interpolation of the first available extracted scanned survival proportions before and after this time point.

When the population *n*_*u*_ is not reported below the PFS and OS Kaplan-Meier curve for certain time points *u*, it can be imputed. First, based on the reported size of the at-risk population at subsequent time points (*n*_*u*+1_), *n*_*u*_ will be estimated according to 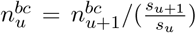. With this ‘backward calculation’ approach we implicitly assume that censoring occurs before the events happen within a time interval. However, this approach is not feasible if there is no information regarding the at-risk population for time intervals beyond the at-risk population reported at a certain time point. In other words, this approach is only feasibly for intervals up to the latest time point for which population is reported. Next, *n*_*u*_ will be estimated according to 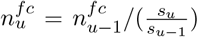. The disadvantage of this ‘forward calculation’ approach is that censoring is ignored and the sample size potentially too large for those timepoints. For intervals where both 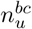 and 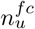 was calculated, the actual estimate for the population at-risk is calculated as: 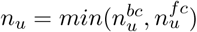 to ensure the sample size is not overestimated. For time points where 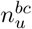 could not be calculated, 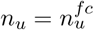.

Based on the subsequent *s*_*u*_ for the four points at each interval (i.e. 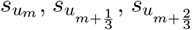, and 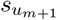), three conditional survival proportions are obtained: 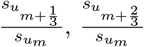, and 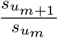. The corresponding sample sizes are defined as 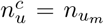. The corresponding observed number of patients who have not yet experienced progression or death are calculated according to 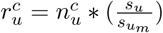.

Applying this algorithm to PFS and OS of each arm *i* of each trial *k*, we get a data set with 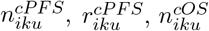 and 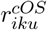.

We set-up the event dataset such that every row represents one time interval with 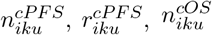, and 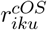 corresponding to 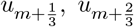, and 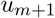. In addition, each row has a variable related to follow-up time 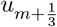, three variables related to 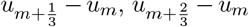, and *u*_*m*+1_ − *u*_*m*_, the study number, and study-arm number within that study.

In additon to the event dataset, we create a study dataset indicating the compared interventions in each study along with the number of study arms. See the supplementary material (Section C.3) for example data structures.

### B States and between-state transition rates

#### B.1 Dynamic transitions - Problem specification

Figure 1 represents a closed dynamic system (*S*_*ik*_(*u*) + *P*_*ik*_(*u*) + *D*_*ik*_(*u*) = 1) whose evolution is determined by a known initial condition at time *u* = 0 and three differential equations:

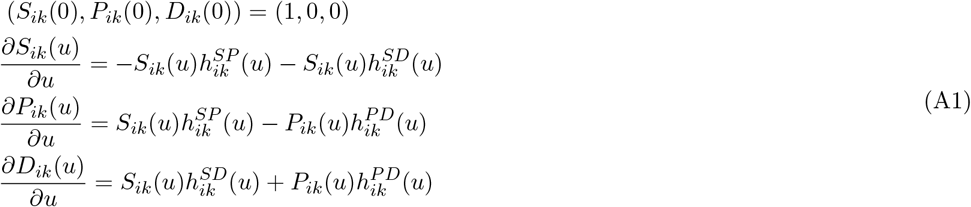

with 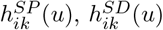, and 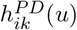 the time-varying hazard rates for the transitions in the figure.

#### B.2 Aproximating arbitrary 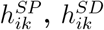, and 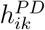

We can approximate arbitrary hazard rate functions with a set of discontinuous constant hazard rates over successive time intervals. We prefer this approximation because the system Equation A1 can be solved analytically when the transition rates are constant using the the eigenvalue method for first-order differential equations. For *u* ∈ *U*_*m*_ Equation A1 become:

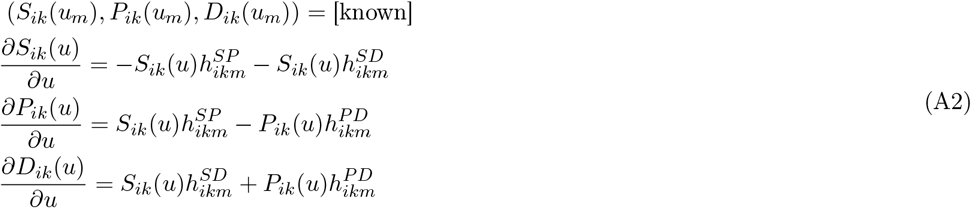

#### B.3 Analytic solutions for *S*_*ik*_(*u*), *P*_*ik*_(*u*), **and** *D*_*ik*_(*u*) **where** *u* ∈ *U*_*m*_

Write the system in Equation A2 in matrix form:

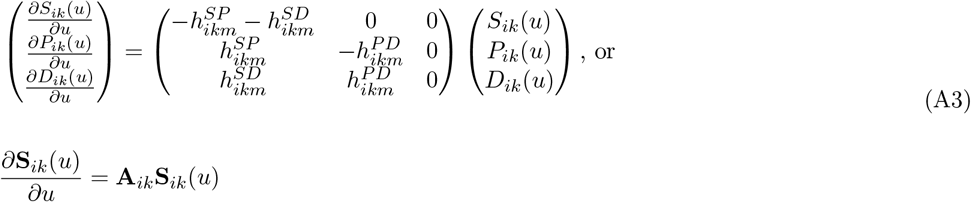

with the obvious notational correspondence between the two equations. For *u* ∈ *U*_*m*_ the system is homogenous and its general solution is the superposition:

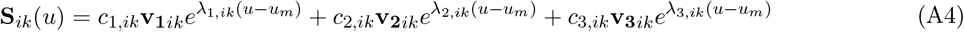

where *λ*_1,*ik*_, *λ*_2,*ik*_, and *λ*_3,*ik*_ are the eigenvalues of the coefficient matrix **A**_*ik*_. **v**_**1***ik*_, **v**_**2***ik*_ and **v**_**3***ik*_ are the corresponding eigenvectors, and *c*_1,*ik*_, *c*_2,*ik*_, *c*_3,*ik*_ scalar constants to be identified from the initial condition in Equation A2. In our case:

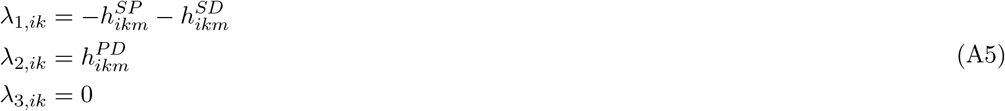

The eigenvectors are:

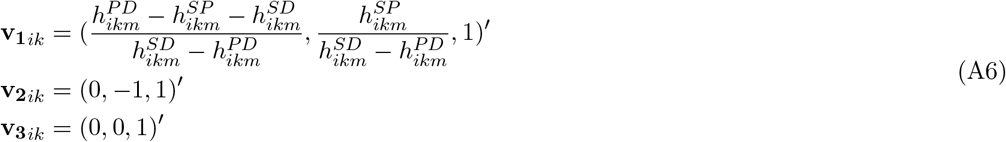

##### B.3.1 Identification of constants in the general solution

The constants *c*_1,*ik*_, *c*_2,*ik*_, *c*_3,*ik*_ are identied from the proportions at the beginning of *U*_*m*_. Setting *u* = *u*_*m*_ in the general solution, and using the initial condition in Equation A1 and Equation A2 we obtain:

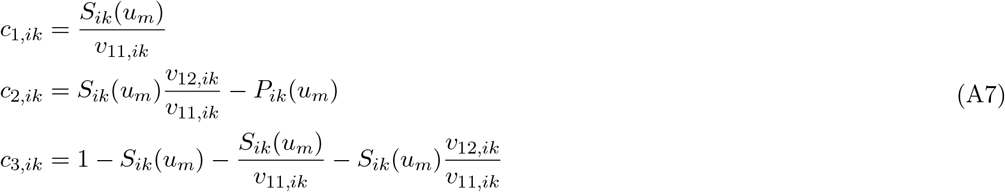

where *v*_*xy,ik*_ is element *x* of eigenvector *y*.

##### B.3.2 Solution for *S*_*ik*_(*u*), *u* ∈ *U*_*m*_

Substituting *c*_1,*ik*_, *c*_2,*ik*_, *c*_3,*ik*_ from Equation A7 in Equation A4 we obtain for *S*_*ik*_(*u*) :

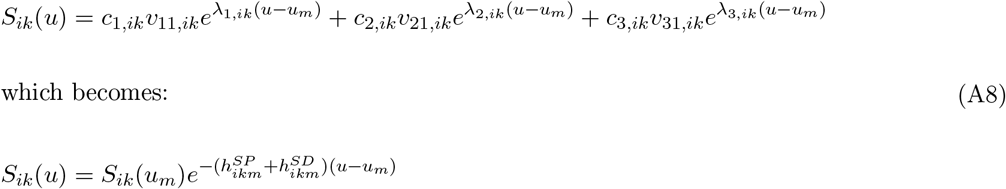

##### B.3.3 Solution for *P*_*ik*_(*u*), *u* ∈ *U*_*m*_

Substituting *c*_1,*ik*_, *c*_2,*ik*_, *c*_3,*ik*_ from Equation A7 in Equation A4 we obtain for *P*_*ik*_(*u*):

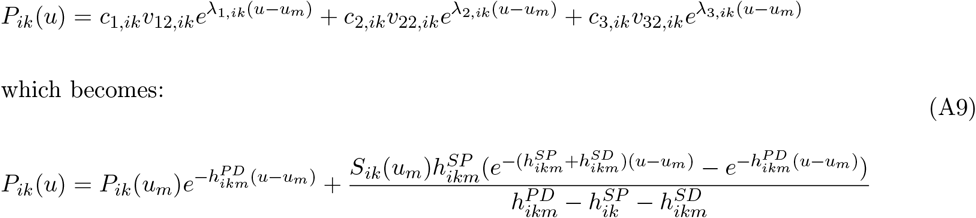

##### B.3.4 Solution for *D*_*ik*_(*u*), *u* ∈ *U*_*m*_

Using Equation A1, Equation A8, and Equation A9 we obtain:

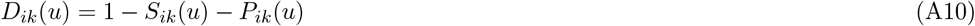

### C Online supplementary material

#### C.1 Kaplan-Meier curves

**Figure A1:**
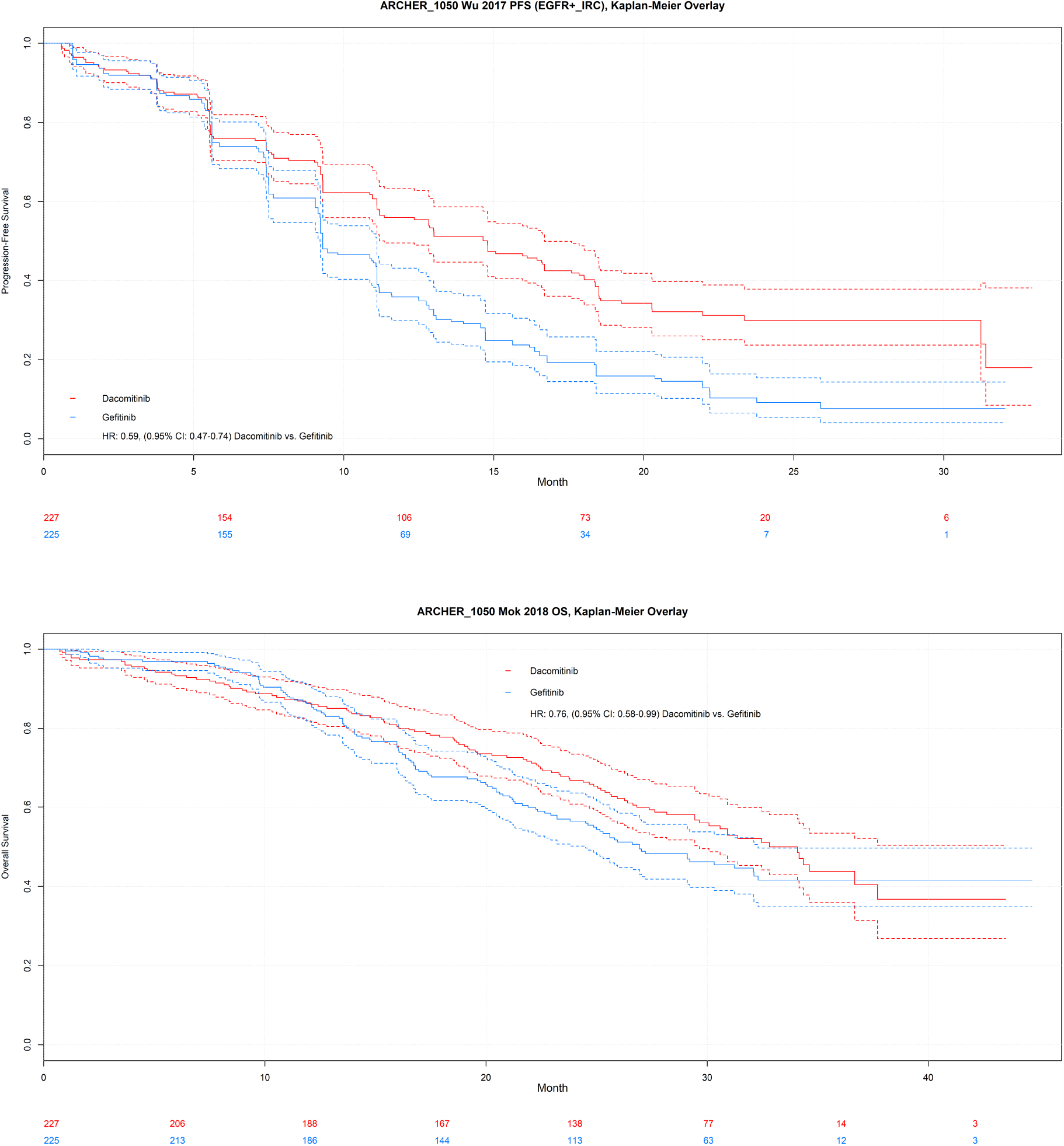
ARCHER-1050, progression-free survival and overall survival.

**Figure A2:**
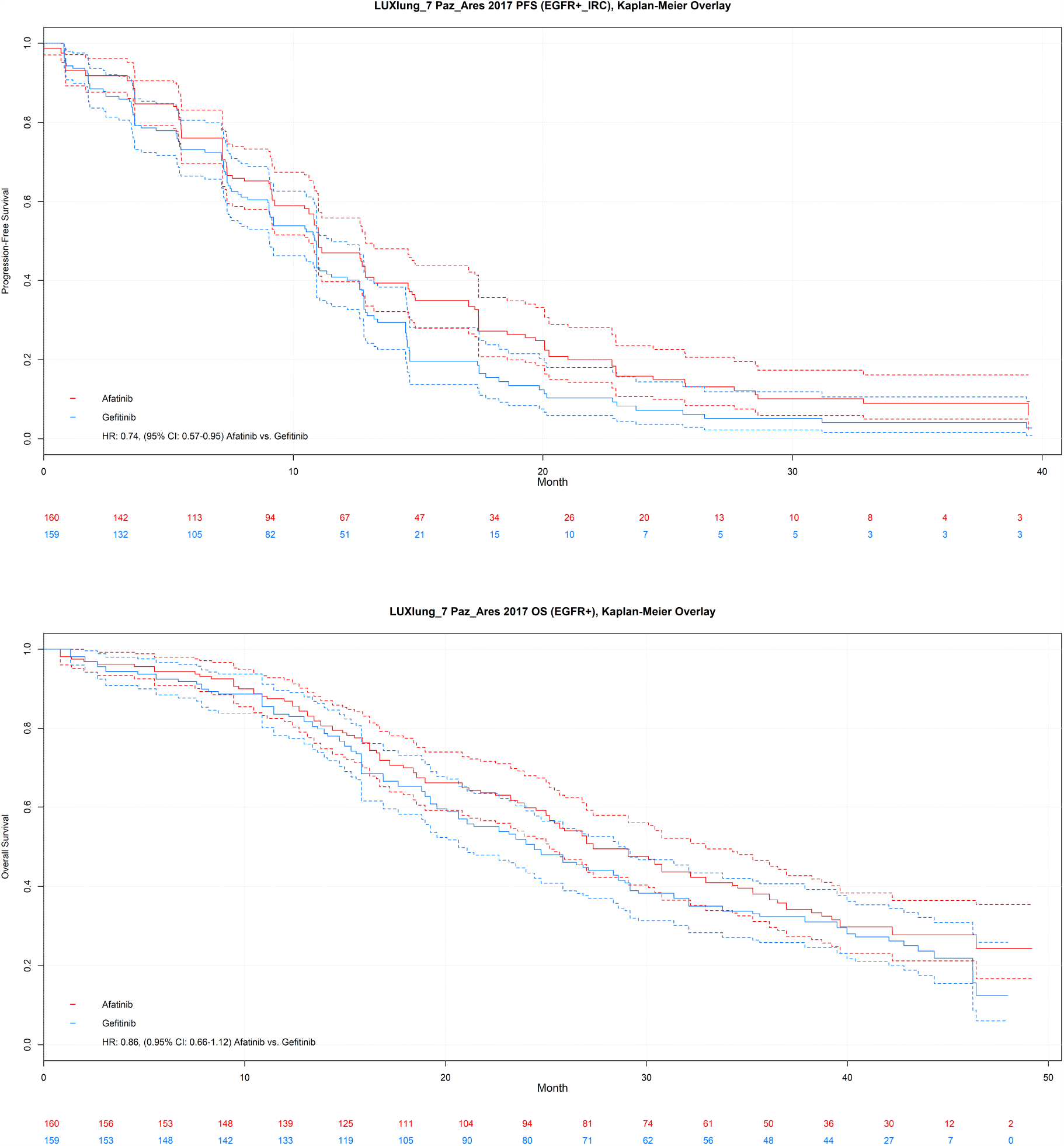
LUX-LUNG 7, progression-free survival and overall survival.

**Figure A3:**
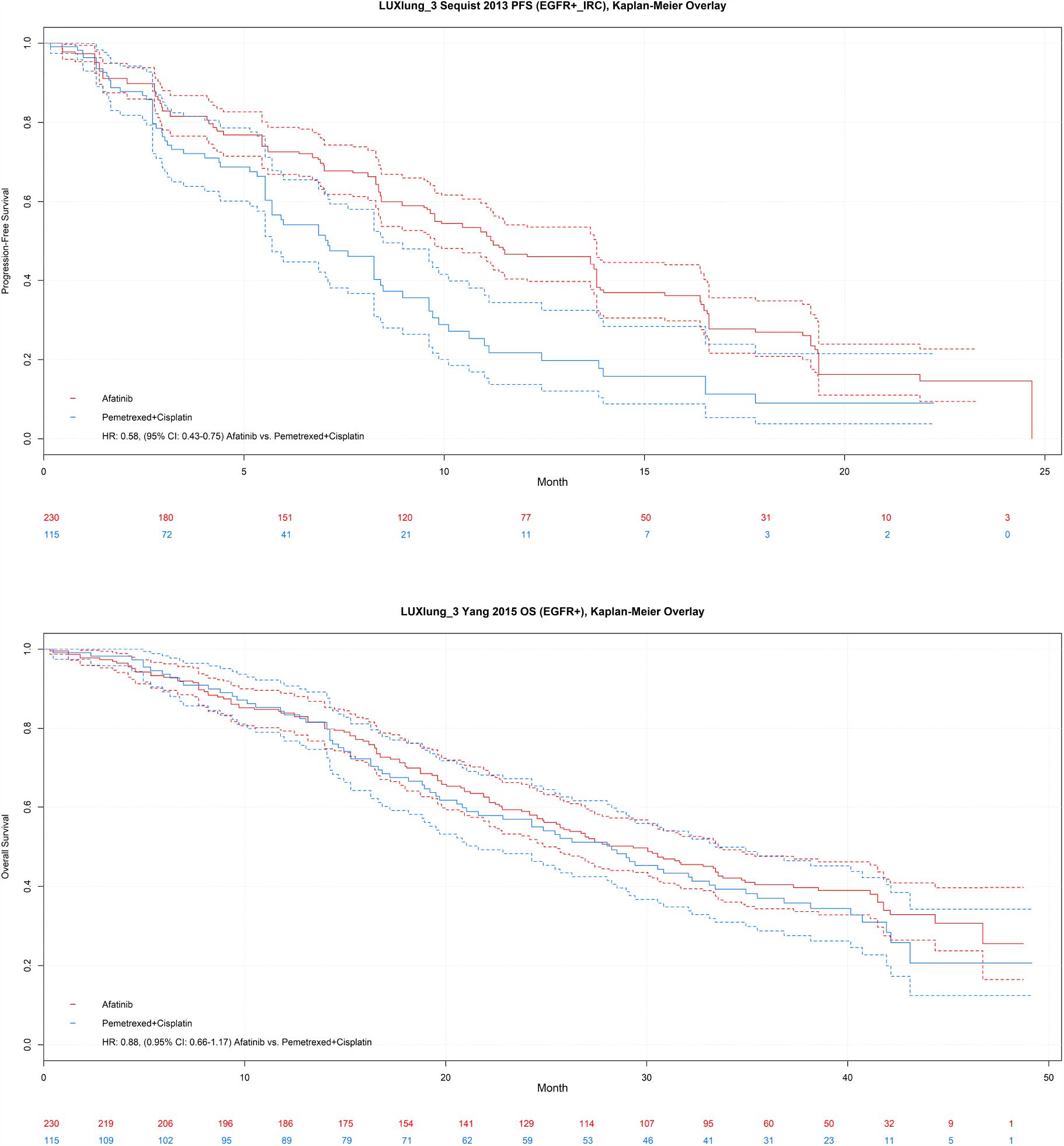
LUX-LUNG 3, progression-free survival and overall survival.

**Figure A4:**
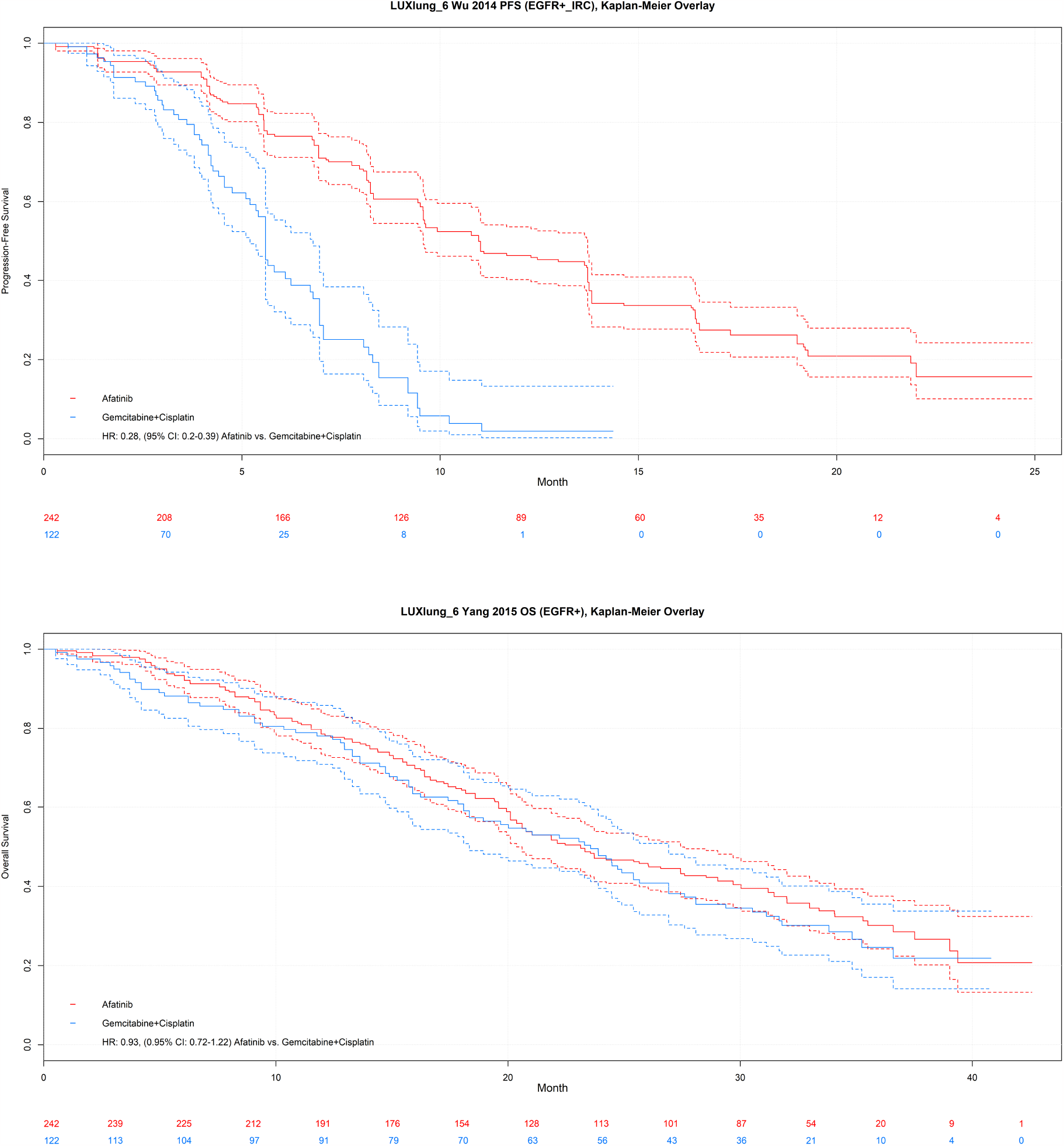
LUX-LUNG 6, progression-free survival and overall survival.

**Figure A5:**
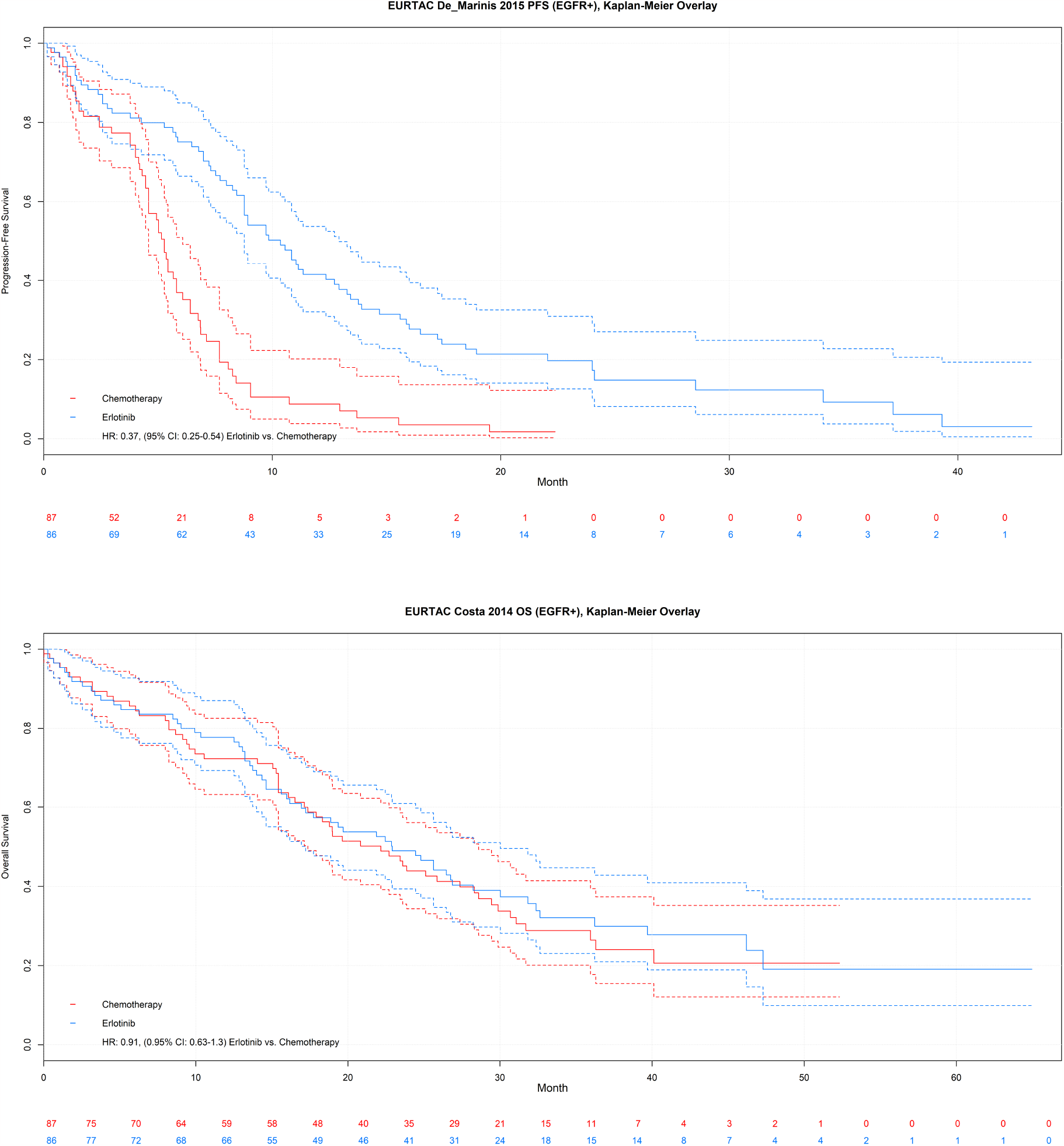
EURTAC, progression-free survival and overall survival.

**Figure A6:**
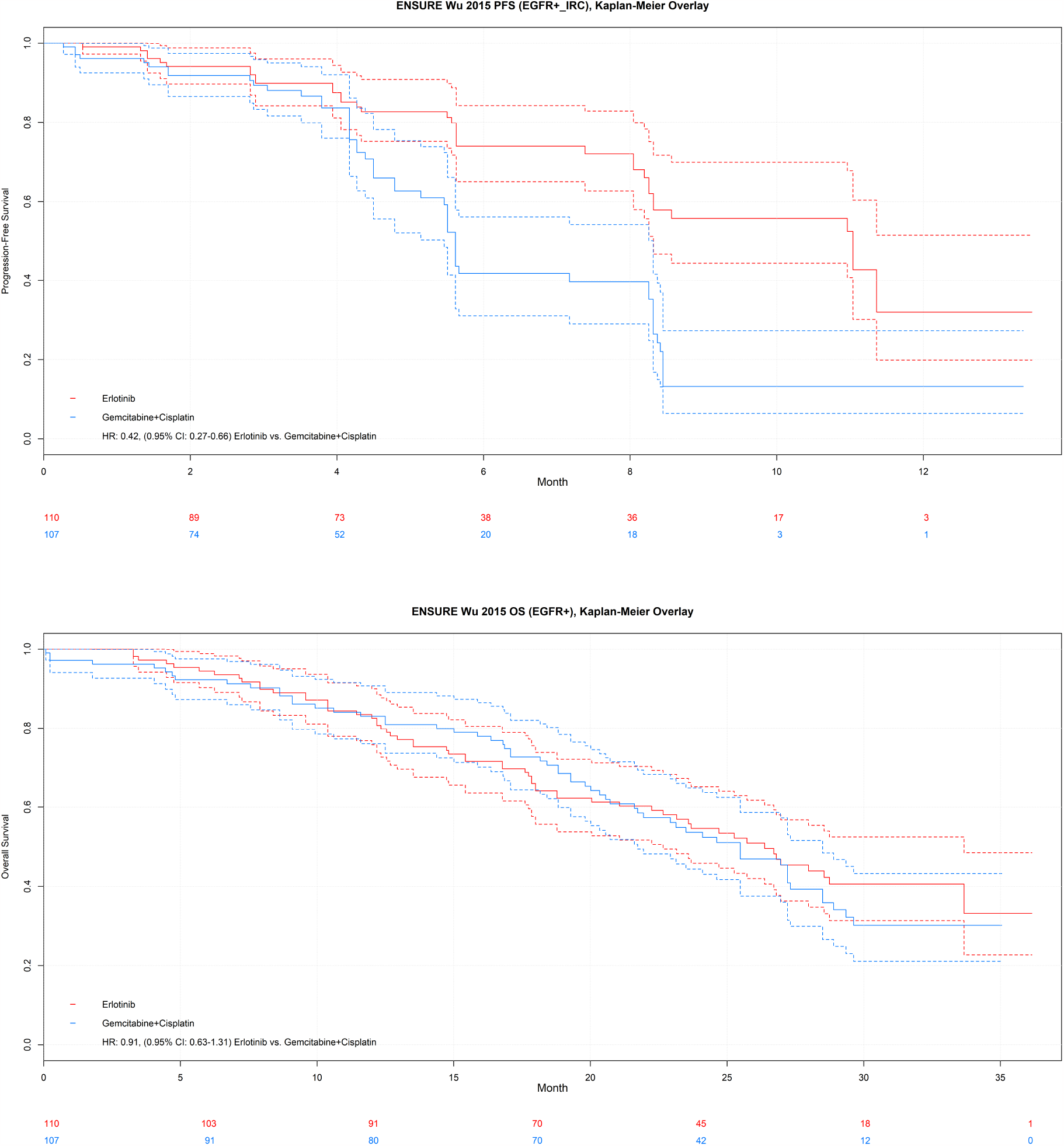
ENSURE, progression-free survival and overall survival.

**Figure A7:**
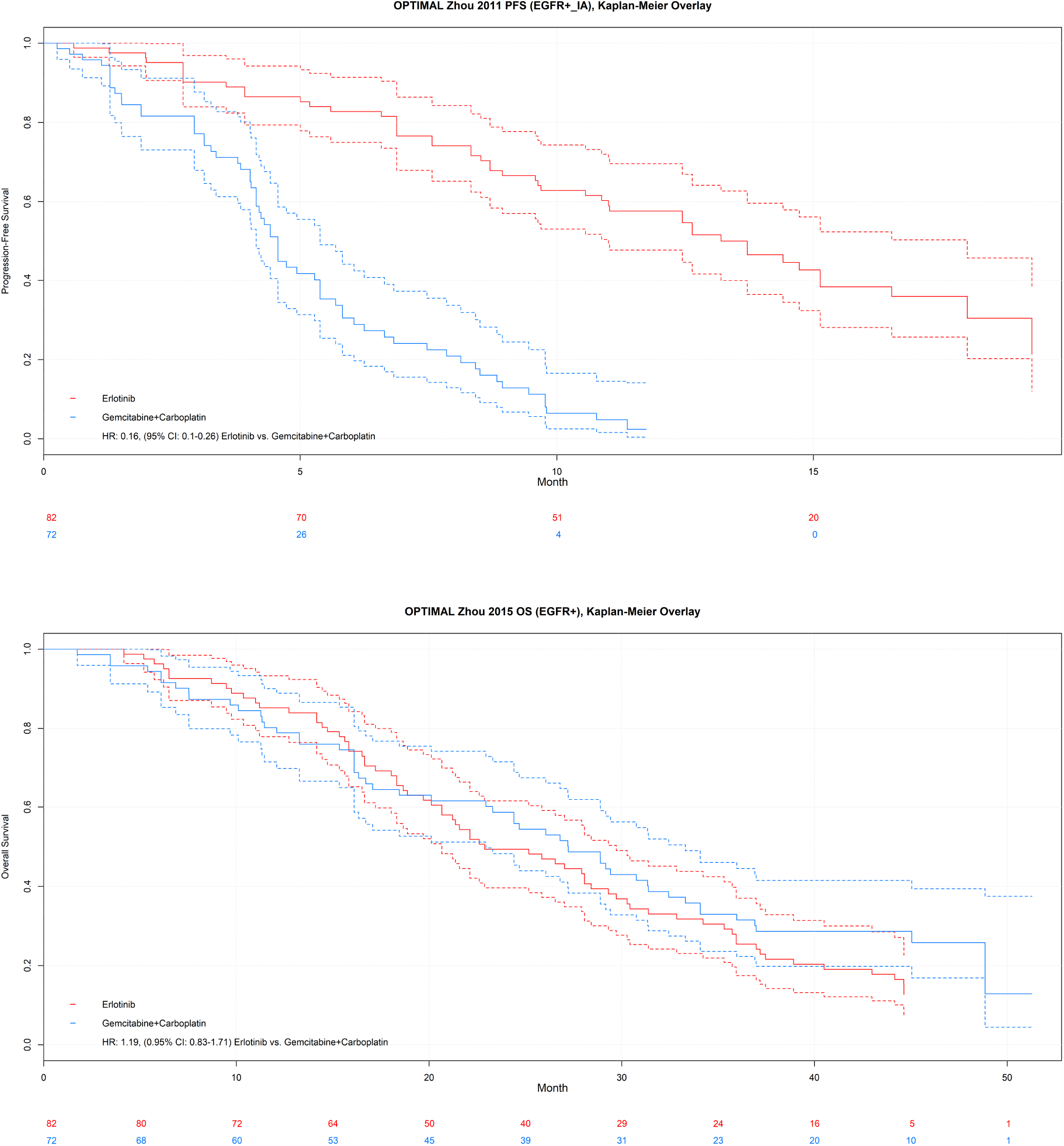
OPTIMAL, progression-free survival and overall survival.

**Figure A8:**
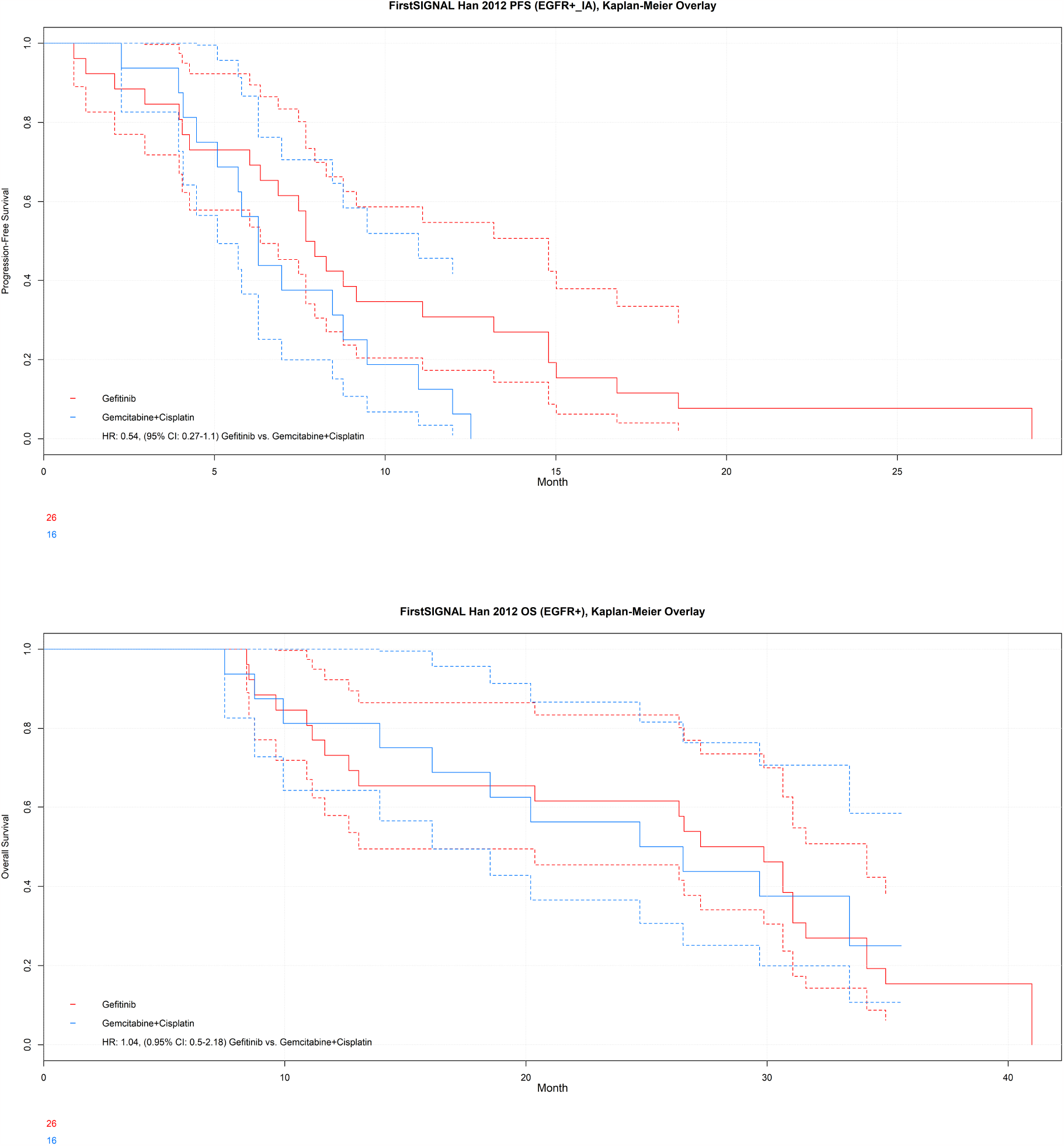
FirstSIGNAL, progression-free survival and overall survival.

**Figure A9:**
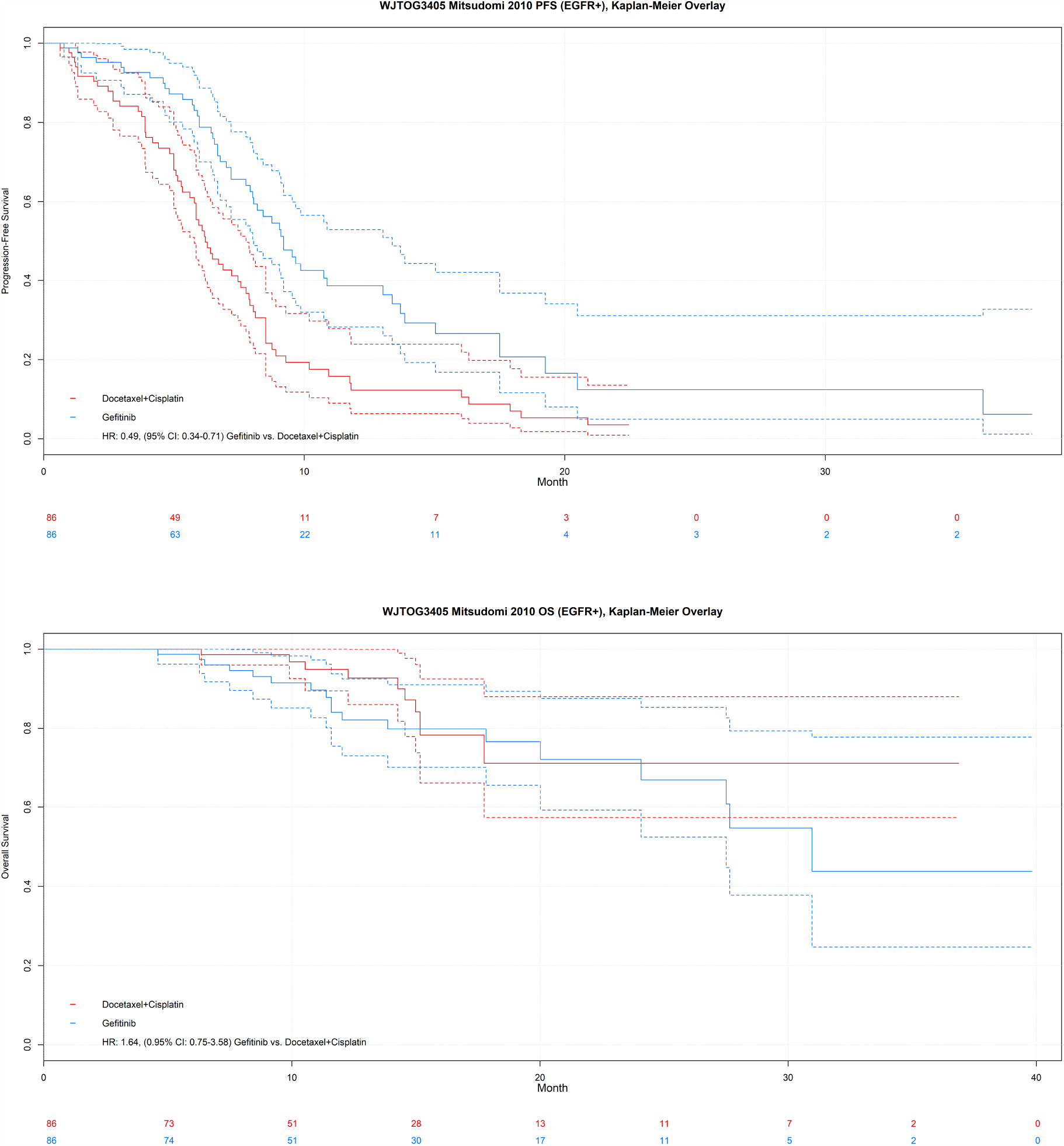
WJTOG3405, progression-free survival and overall survival.

**Figure A10:**
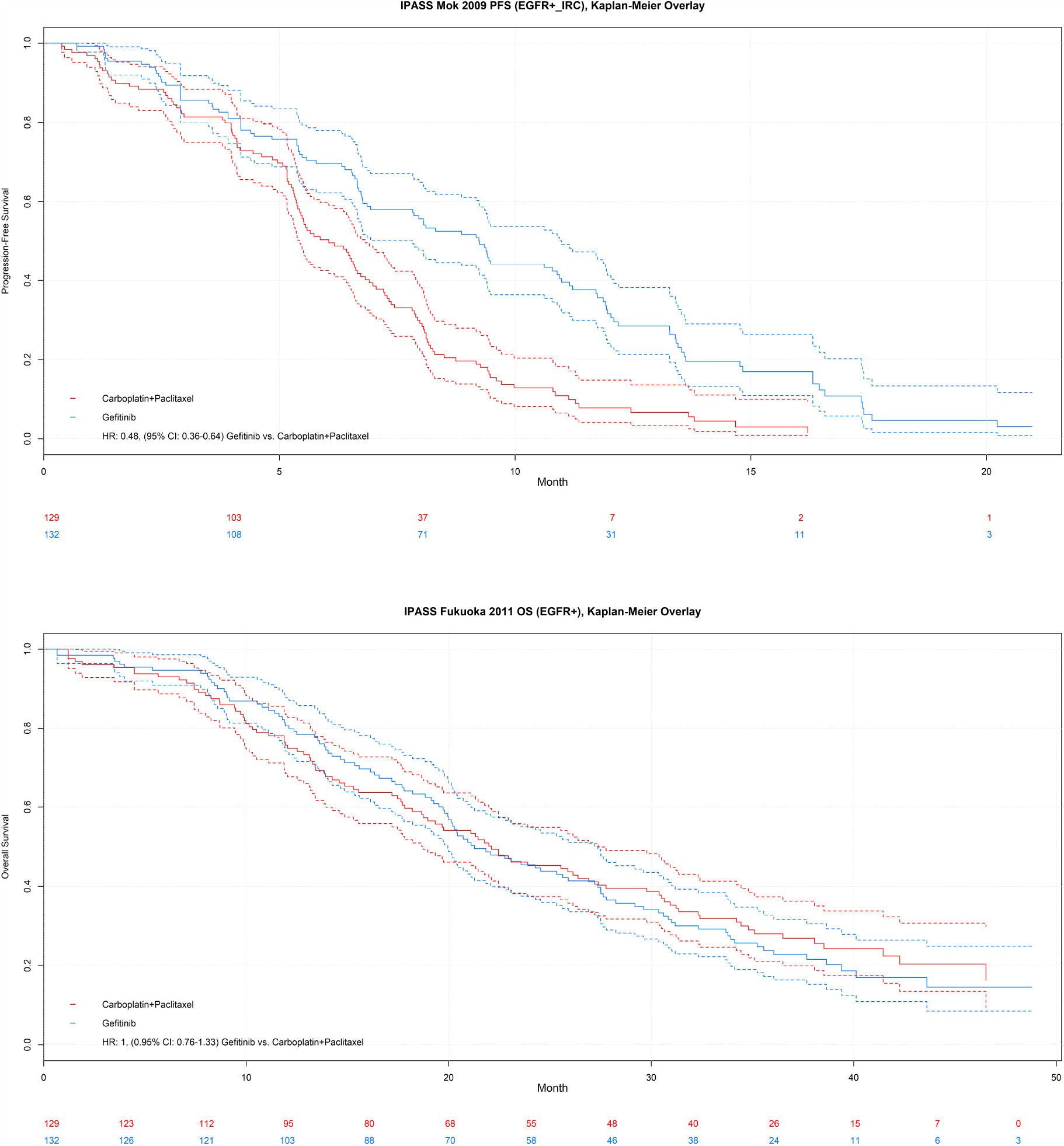
IPASS, progression-free survival and overall survival.

**Figure A11:**
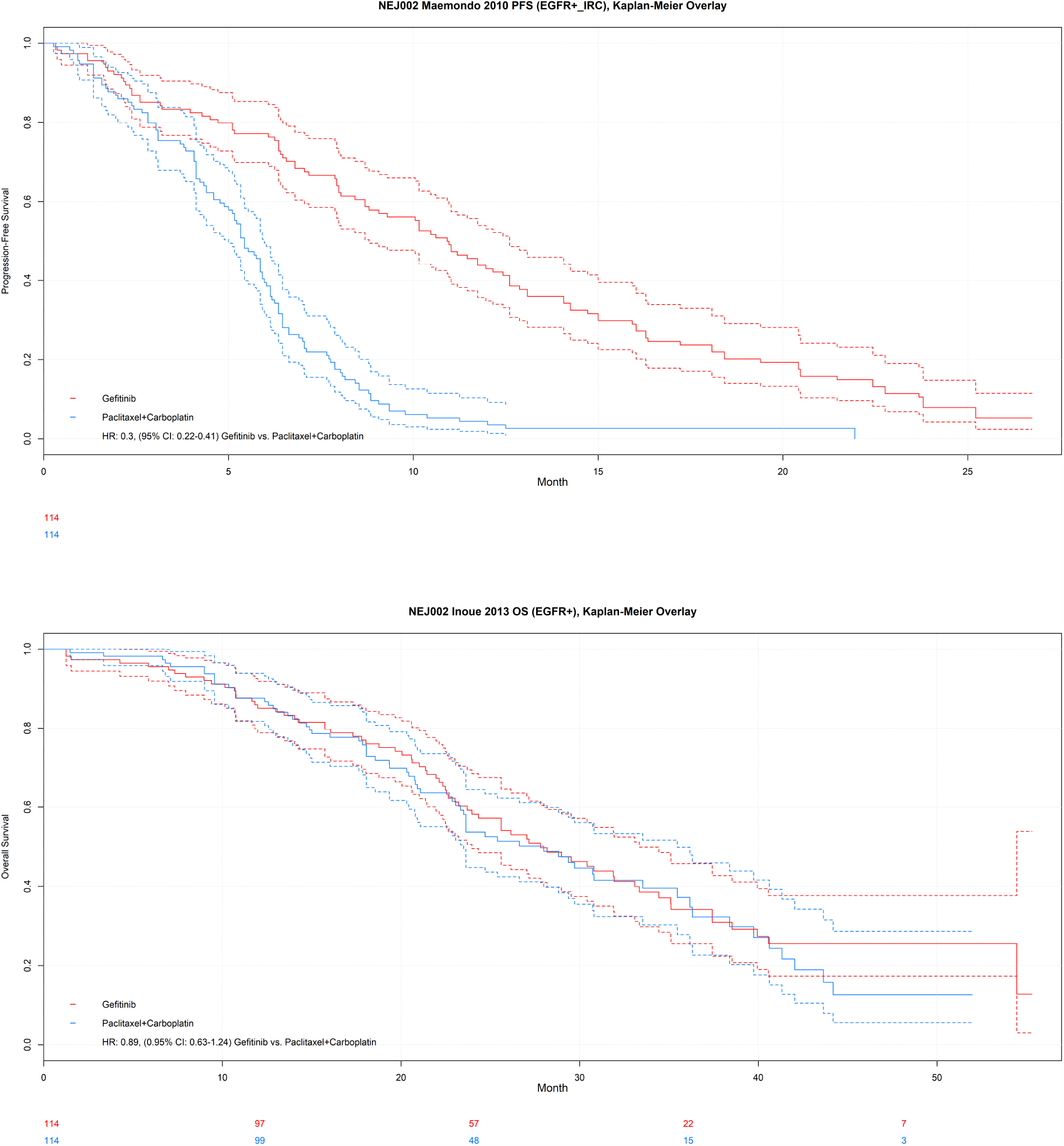
NEJ002, progression-free survival and overall survival.

**Figure A12:**
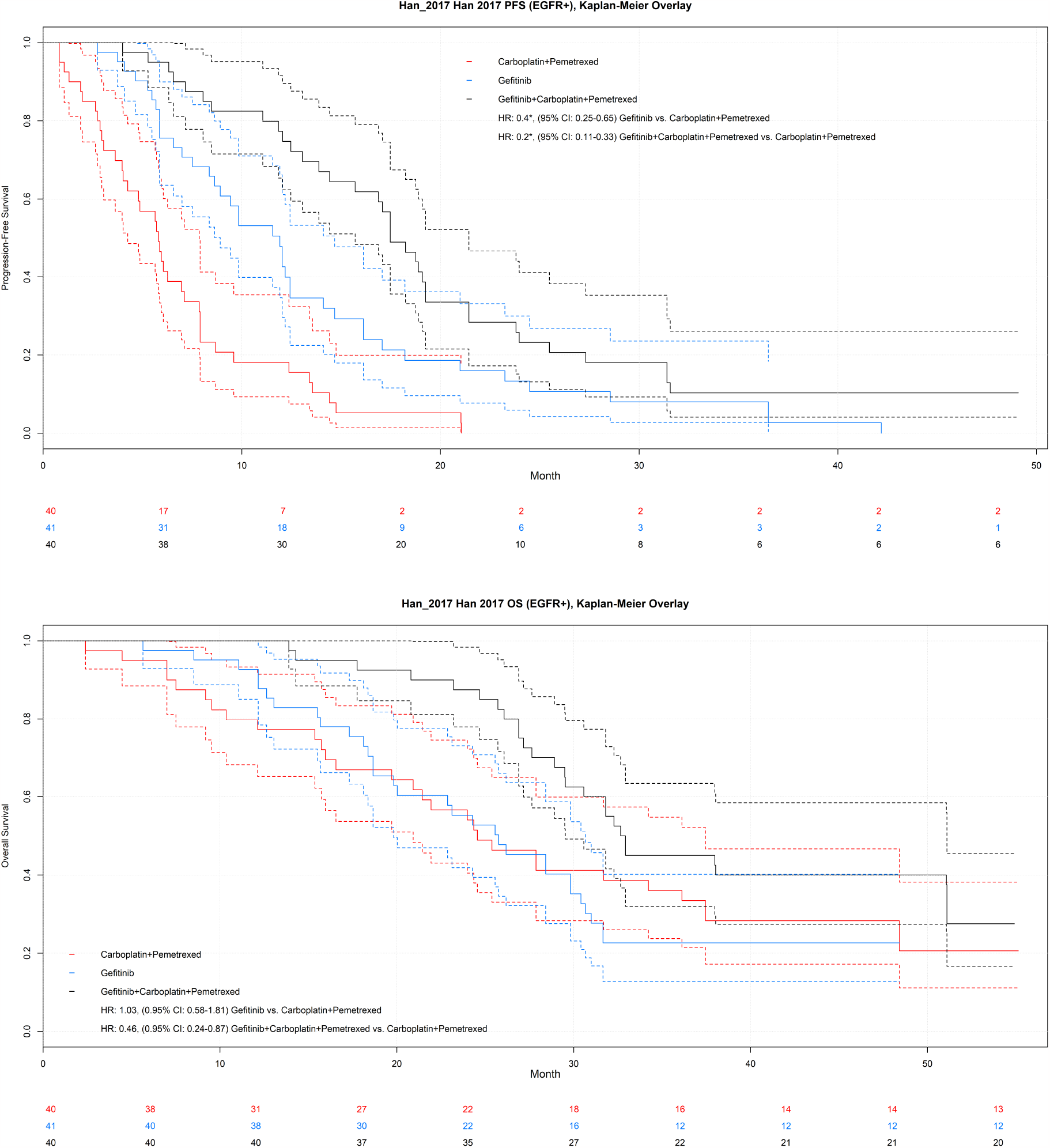
Han 2017, progression-free survival and overall survival.

**Figure A13:**
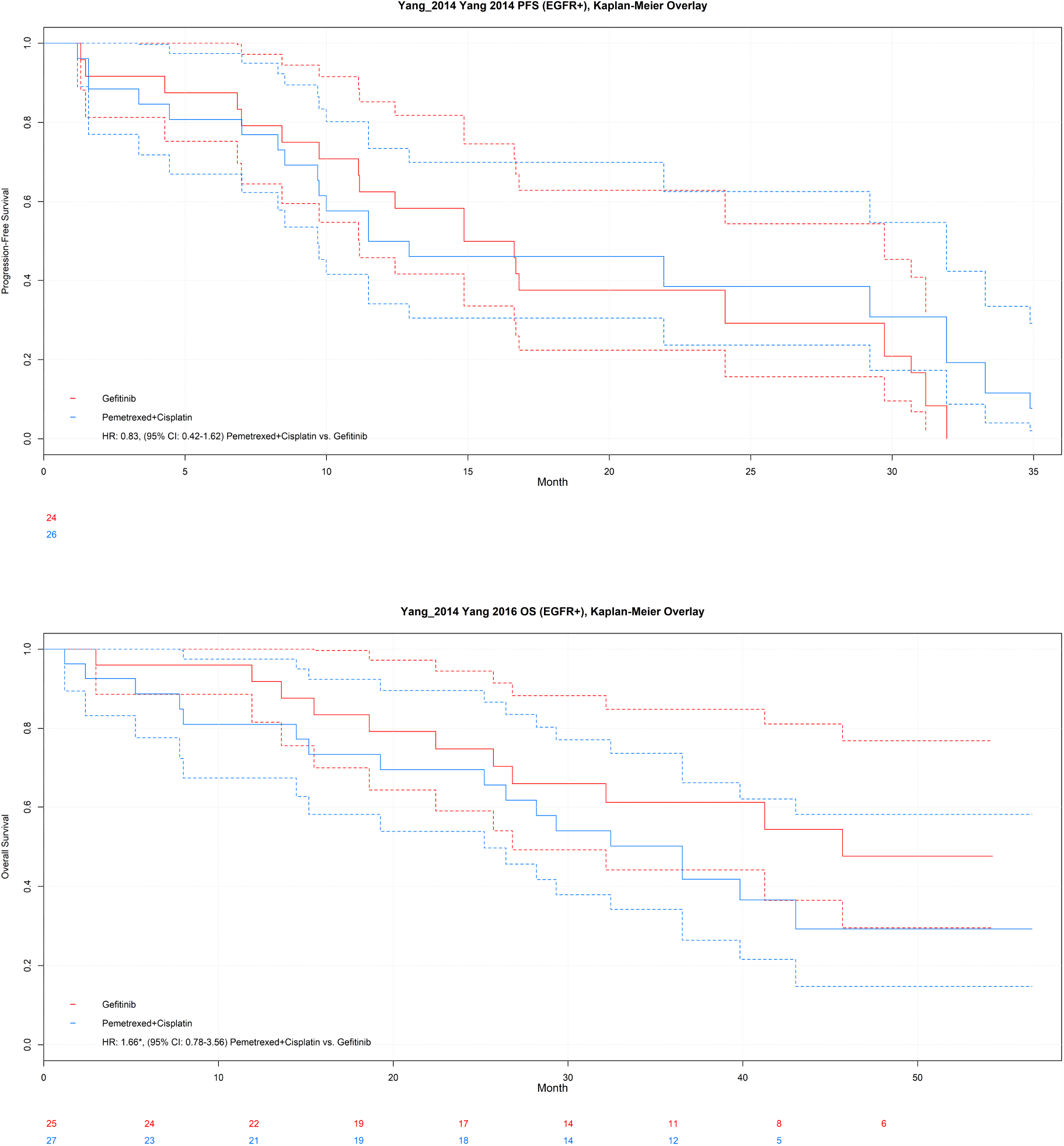
Yang 2014 and Yang 2016, progression-free survival and overall survival.

#### C.2 JAGS code random effects NMA model

Example JAGS code corresponding to model *SP Weibull RE2; SD exponential; PD Weibull FE1(scale)* has been presented here.

~~~
model{
# Likelihood for conditional survival probabilities
for (j in 1:Nd){
  r_cond_pfs1[j]∼dbinom(cond_pfs[j,1], n_cond_pfs1[j])
  r_cond_pfs2[j]∼dbinom(cond_pfs[j,2], n_cond_pfs2[j])
  r_cond_pfs3[j]∼dbinom(cond_pfs[j,3], n_cond_pfs3[j])
  r_cond_os1[j]∼dbinom(cond_os[j,1], n_cond_os1[j])
  r_cond_os2[j]∼dbinom(cond_os[j,2], n_cond_os2[j])
  r_cond_os3[j]∼dbinom(cond_os[j,3], n_cond_os3[j])
}
# transformation of conditional survival probabilities to survival probabilities
p4[1]<-1
p5[1]<-0
for (j in 2:Nd){
 p4[j]<-equals(a[j]-a[j-1],0)*p[(j-1),4] + (1-equals(a[j]-a[j-1],0))*1
 p5[j]<-equals(a[j]-a[j-1],0)*p[(j-1),5] + (1-equals(a[j]-a[j-1],0))*0
}
for (j in 1:Nd){
 cond_pfs[j,1]<-p[j,1]/p4[j]
 cond_pfs[j,2]<-p[j,4]/p4[j]
 cond_pfs[j,3]<-p[j,7]/p4[j]
 cond_os[j,1]<-(p[j,1]+p[j,2])/(p4[j]+p5[j])
 cond_os[j,2]<-(p[j,4]+p[j,5])/(p4[j]+p5[j])
 cond_os[j,3]<-(p[j,7]+p[j,8])/(p4[j]+p5[j])
 # transformation of survival probabilities in time-varying hazards
 # for transitions between health states
 p[j,1]<- p4[j]*exp(-(h.sd[j]+h.sp[j])*dt[j,1])
 p[j,2]<- p5[j]*exp(-h.pd[j]*dt[j,1])+p4[j]*h.sp[j]*(exp(-(h.sd[j]+h.sp[j])*dt[j,1])
                -exp(-h.pd[j]*dt[j,1]))/(h.pd[j]-h.sp[j]-h.sd[j])
 p[j,3]<-1-(p[j,1]+p[j,2])
 p[j,4]<- p4[j]*exp(-(h.sd[j]+h.sp[j])*dt[j,2])
 p[j,5]<- p5[j]*exp(-h.pd[j]*dt[j,2])+p4[j]*h.sp[j]*(exp(-(h.sd[j]+h.sp[j])*dt[j,2])
                -exp(-h.pd[j]*dt[j,2]))/(h.pd[j]-h.sp[j]-h.sd[j])
 p[j,6]<-1-(p[j,4]+p[j,5])
 p[j,7]<- p4[j]*exp(-(h.sd[j]+h.sp[j])*dt[j,3])
 p[j,8]<- p5[j]*exp(-h.pd[j]*dt[j,3])+p4[j]*h.sp[j]*(exp(-(h.sd[j]+h.sp[j])*dt[j,3])
               -exp(-h.pd[j]*dt[j,3]))/(h.pd[j]-h.sp[j]-h.sd[j])
 p[j,9]<-1-(p[j,7]+p[j,8])
 # constraint to avoid “closing off” SD or PD path
 constraint.sd[j] ∼ dinterval(h.sd[j], EPSILON[1]) # h.sd[j] > EPSILON[1] (e.g. >0.001)
 constraint.pd[j] ∼ dinterval(h.pd[j], EPSILON[2]) # h.pd[j] > EPSILON[2] (e.g. >0.001)
 # decribe hazards as a function of time
 log(h.sp[j])<- alpha[s[j],a[j],1]+alpha[s[j],a[j],2]*timetrans1[j]
 log(h.sd[j])<- alpha[s[j],a[j],3]
 log(h.pd[j])<- alpha[s[j],a[j],4]+alpha[s[j],a[j],5]*timetrans1[j]
}
# random effects model
for (i in 1:Ns){
  for (k in 1:na[i]){
    alpha[i,k,1]<-mu[i,1]+delta[i,k]
    alpha[i,k,2]<-mu[i,2]+d[t[i,k],2]-d[t[i,1],2]
    alpha[i,k,3]<-mu[i,3]
    alpha[i,k,4]<-mu[i,4]+d[t[i,k],3]-d[t[i,1],3]
    alpha[i,k,5]<-mu[i,5]
}
w[i,1]<-0
delta[i,1]<-0
for (k in 2:na[i]){
  delta[i,k]∼dnorm(md[i,k],taud[i,k])
  md[i,k]<-d[t[i,k],1]-d[t[i,1],1] +sw[i,k]
  w[i,k] <- (delta[i,k] - d[t[i,k],1] + d[t[i,1],1])
  sw[i,k] <- sum(w[i,1:(k-1)])/(k-1)
  taud[i,k] <- tau *2*(k-1)/k
 }
}
# priors
for (i in 1:Ns){
  mu[i,1:5] ∼ dmnorm(prior_mean_mu[1:5],prior_varcov_mu[,])
}
d[1,1]<-0
d[1,2]<-0
d[1,3]<-0
for (T in 2:Nt){
  d[T,1:3] ∼ dmnorm(prior_mean_d[1:3],prior_varcov_d[,])
}
sd∼dunif(0,2)
tau<-1/(sd*sd)
# output
for (T in 2:Nt){
  for (u in 1:48){
    log(HR.SP[1,T,u])<-(d[T,1]-d[1,1])+(d[T,2]-d[1,2])*log(u)
    log(HR.SD[1,T,u])<-0
    log(HR.PD[1,T,u])<-(d[T,3]-d[1,3])
  }
}
Mu_mean[1]<- -3.563312141
Mu_mean[2]<- 0.486802196
Mu_mean[3]<- -4.580999291
Mu_mean[4]<- -3.488028822
Mu_mean[5]<- 0.174942184
for (T in 1:Nt){
  Alpha1[T]<-Mu_mean[1]+d[T,1]
  Alpha2[T]<-Mu_mean[2]+d[T,2]
  Alpha3[T]<-Mu_mean[3]
  Alpha4[T]<-Mu_mean[4]+d[T,3]
  Alpha5[T]<-Mu_mean[5]
}
for (T in 1:Nt){
  for (u in 1:48){
    log(HAZARD.SP[T,u])<-(Alpha1[T])+(Alpha2[T])*log(u)
    log(HAZARD.SD[T,u])<-(Alpha3[T])
    log(HAZARD.PD[T,u])<-(Alpha4[T])+(Alpha5[T])*log(u)
   }}
for (T in 1:Nt){
  P.S[T,1]<-1*exp(-(HAZARD.SD[T,1]+HAZARD.SP[T,1]))
  P.P[T,1]<-0*exp(-HAZARD.PD[T,1])+1*HAZARD.SP[T,1]*(exp(-(HAZARD.SD[T,1]+HAZARD.SP[T,1]))
                -exp(-HAZARD.PD[T,1]))/(HAZARD.PD[T,1]-HAZARD.SP[T,1]-HAZARD.SD[T,1])
  PFS[T,1]<-P.S[T,1]
  OS[T,1]<-P.S[T,1]+P.P[T,1]
  for (u in 2:48){
    P.S[T,u]<-P.S[T,(u-1)]*exp(-(HAZARD.SD[T,u]+HAZARD.SP[T,u]))
    P.P[T,u]<-P.P[T,(u-1)]*exp(-HAZARD.PD[T,u])+P.S[T,(u-1)]*HAZARD.SP[T,u]
              *(exp(-(HAZARD.SD[T,u]+HAZARD.SP[T,u]))
              -exp(-HAZARD.PD[T,u]))/(HAZARD.PD[T,u]-HAZARD.SP[T,u]-HAZARD.SD[T,u])
    PFS[T,u]<-P.S[T,u]
    OS[T,u]<-P.S[T,u]+P.P[T,u]
   }
 }
}
~~~

#### C.3 Data structures

**Table A1:** Data structure for the study level information: treatment by study-arm and number of study-arms.

| **t[,1]** | **t[,2]** | **na[]** | **Study** | **Treatment arm 1** | **Treatment arm 2** |
| --- | --- | --- | --- | --- | --- |
| 1 | 5 | 2 | Yang-2014 | Gefitinib | PBDC |
| 1 | 5 | 2 | WJTOG3405 | Gefitinib | PBDC |
| 2 | 5 | 2 | OPTIMAL | Erlotinib | PBDC |
| 1 | 5 | 2 | NEJ002 | Gefitinib | PBDC |
| 1 | 3 | 2 | LUXlung_7 | Gefitinib | Afatinib |
| 3 | 5 | 2 | LUXlung_6 | Afatinib | PBDC |
| 3 | 5 | 2 | LUXlung_3 | Afatinib | PBDC |
| 1 | 5 | 2 | IPASS | Gefitinib | PBDC |
| 1 | 5 | 2 | Han_2017 | Gefitinib | PBDC |
| 1 | 5 | 2 | FirstSIGNAL | Gefitinib | PBDC |
| 2 | 5 | 2 | EURTAC | Erlotinib | PBDC |
| 2 | 5 | 2 | ENSURE | Erlotinib | PBDC |
| 1 | 4 | 2 | ARCHER_1050 | Gefitinib | Dacomitinib |

t[,1]: treatment in arm 1
t[,2]: treatment in arm 2
na[]: number of arms in study
PBDC: platinum-based doublet chemotherapy

**Table A2:** Data structure regarding the conditional PFS and OS probablities at the three time points per interval over time.

| s[] | a[] | time | dt[,1] | dt[,2] | dt[,3] | r_cond_pfs1[] | n_cond_pfs1[] | r_cond_pfs2[] | n_cond_pfs2[] | r_cond_pfs3[] | n_cond_pfs3[] | r_cond_os1[] | n_cond_os1[] | r_cond_os2[] | n_cond_os2[] | r_cond_os3[] | n_cond_os3[] |
| --- | --- | --- | --- | --- | --- | --- | --- | --- | --- | --- | --- | --- | --- | --- | --- | --- | --- |
| 1 | 1 | 1 | 1 | 2 | 3 | 23 | 24 | 22 | 24 | 22 | 24 | 25 | 25 | 25 | 25 | 24 | 25 |
| 1 | 1 | 4 | 1 | 2 | 3 | 22 | 22 | 21 | 22 | 20 | 22 | 24 | 24 | 24 | 24 | 24 | 24 |
| 1 | 1 | 7 | 1 | 2 | 3 | 20 | 21 | 19 | 21 | 18 | 21 | 24 | 24 | 24 | 24 | 24 | 24 |
| 1 | 1 | 10 | 1 | 2 | 3 | 18 | 19 | 18 | 19 | 16 | 19 | 22 | 22 | 21 | 22 | 21 | 22 |
| 1 | 1 | 13 | 1 | 2 | 3 | 15 | 16 | 15 | 16 | 13 | 16 | 21 | 22 | 21 | 22 | 20 | 22 |
| 1 | 1 | 16 | 1 | 2 | 3 | 14 | 14 | 10 | 14 | 10 | 14 | 19 | 19 | 19 | 19 | 19 | 19 |
| 1 | 1 | 19 | 1 | 2 | 3 | 10 | 10 | 10 | 10 | 10 | 10 | 18 | 19 | 18 | 19 | 18 | 19 |
| 1 | 1 | 22 | 1 | 2 | 3 | 10 | 10 | 10 | 10 | 10 | 10 | 18 | 18 | 17 | 18 | 17 | 18 |
| 1 | 1 | 25 | 1 | 2 | 3 | 9 | 10 | 9 | 10 | 9 | 10 | 16 | 17 | 16 | 17 | 15 | 17 |
| 1 | 1 | 28 | 1 | 2 | 3 | 9 | 9 | 9 | 9 | 7 | 9 | 14 | 14 | 14 | 14 | 14 | 14 |
| 1 | 1 | 31 | 1 | 2 | 3 |  | 7 |  | 7 |  | 7 | 14 | 14 | 14 | 14 | 13 | 14 |
| 1 | 1 | 34 | 1 | 2 | 3 |  |  |  |  |  |  | 11 | 11 | 11 | 11 | 11 | 11 |
| 1 | 1 | 37 | 1 | 2 | 3 |  |  |  |  |  |  | 11 | 11 | 11 | 11 | 11 | 11 |
| 1 | 1 | 40 | 1 | 2 | 3 |  |  |  |  |  |  | 9 | 9 | 9 | 9 | 8 | 9 |
| 1 | 1 | 43 | 1 | 2 | 3 |  |  |  |  |  |  | 8 | 8 | 8 | 8 | 8 | 8 |
| 1 | 2 | 1 | 1 | 2 | 3 | 26 | 26 | 23 | 26 | 23 | 26 | 27 | 27 | 26 | 27 | 25 | 27 |
| 1 | 2 | 4 | 1 | 2 | 3 | 22 | 23 | 21 | 23 | 21 | 23 | 24 | 24 | 24 | 24 | 23 | 24 |
| 1 | 2 | 7 | 1 | 2 | 3 | 20 | 21 | 20 | 21 | 18 | 21 | 23 | 23 | 21 | 23 | 21 | 23 |
| 1 | 2 | 10 | 1 | 2 | 3 | 15 | 18 | 15 | 18 | 14 | 18 | 21 | 21 | 21 | 21 | 21 | 21 |
| 1 | 2 | 13 | 1 | 2 | 3 | 13 | 14 | 13 | 14 | 13 | 14 | 21 | 21 | 20 | 21 | 20 | 21 |
| 1 | 2 | 16 | 1 | 2 | 3 | 13 | 13 | 13 | 13 | 13 | 13 | 19 | 20 | 19 | 20 | 19 | 20 |
| 1 | 2 | 19 | 1 | 2 | 3 | 13 | 13 | 13 | 13 | 13 | 13 | 18 | 19 | 18 | 19 | 18 | 19 |
| 1 | 2 | 22 | 1 | 2 | 3 | 11 | 13 | 11 | 13 | 11 | 13 | 18 | 18 | 18 | 18 | 18 | 18 |
| 1 | 2 | 25 | 1 | 2 | 3 | 11 | 11 | 11 | 11 | 11 | 11 | 18 | 18 | 16 | 18 | 16 | 18 |
| 1 | 2 | 28 | 1 | 2 | 3 | 11 | 11 | 11 | 11 | 8 | 11 | 14 | 15 | 14 | 15 | 13 | 15 |
| 1 | 2 | 31 | 1 | 2 | 3 | 8 | 8 | 5 | 8 | 5 | 8 | 14 | 14 | 14 | 14 | 13 | 14 |
| 1 | 2 | 34 | 1 | 2 | 3 |  | 5 |  | 5 |  | 5 | 12 | 12 | 12 | 12 | 12 | 12 |
| 1 | 2 | 37 | 1 | 2 | 3 |  |  |  |  |  |  | 11 | 12 | 11 | 12 | 11 | 12 |
| 1 | 2 | 40 | 1 | 2 | 3 |  |  |  |  |  |  | 5 | 6 | 5 | 6 | 5 | 6 |
| ... | ... | ... | ... | ... | ... | ... | ... | ... | ... | ... | ... | ... | ... | ... | ... | ... | ... |
| ... | ... | ... | ... | ... | ... | ... | ... | ... | ... | ... | ... | ... | ... | ... | ... | ... | ... |
| ... | ... | ... | ... | ... | ... | ... | ... | ... | ... | ... | ... | ... | ... | ... | ... | ... | ... |
| 13 | 2 | 1 | 1 | 2 | 3 | 219 | 227 | 212 | 227 | 209 | 227 | 224 | 227 | 221 | 227 | 221 | 227 |
| 13 | 2 | 4 | 1 | 2 | 3 | 177 | 186 | 176 | 186 | 154 | 186 | 211 | 215 | 208 | 215 | 206 | 215 |
| 13 | 2 | 7 | 1 | 2 | 3 | 153 | 154 | 144 | 154 | 143 | 154 | 204 | 206 | 202 | 206 | 198 | 206 |
| 13 | 2 | 10 | 1 | 2 | 3 | 118 | 133 | 114 | 133 | 106 | 133 | 193 | 195 | 190 | 195 | 187 | 195 |
| 13 | 2 | 13 | 1 | 2 | 3 | 99 | 106 | 97 | 106 | 89 | 106 | 186 | 188 | 183 | 188 | 181 | 188 |
| 13 | 2 | 16 | 1 | 2 | 3 | 82 | 84 | 76 | 84 | 73 | 84 | 173 | 178 | 170 | 178 | 167 | 178 |
| 13 | 2 | 19 | 1 | 2 | 3 | 62 | 73 | 61 | 73 | 58 | 73 | 162 | 167 | 157 | 167 | 156 | 167 |
| 13 | 2 | 22 | 1 | 2 | 3 | 21 | 22 | 21 | 22 | 21 | 22 | 147 | 150 | 142 | 150 | 138 | 150 |
| 13 | 2 | 25 | 1 | 2 | 3 | 20 | 20 | 20 | 20 | 20 | 20 | 134 | 138 | 128 | 138 | 123 | 138 |
| 13 | 2 | 28 | 1 | 2 | 3 | 6 | 6 | 6 | 6 | 6 | 6 | 80 | 82 | 79 | 82 | 77 | 82 |
| 13 | 2 | 31 | 1 | 2 | 3 | 6 | 6 |  | 6 |  | 6 | 73 | 77 | 72 | 77 | 69 | 77 |
| 13 | 2 | 34 | 1 | 2 | 3 |  |  |  |  |  |  | 16 | 16 | 14 | 16 | 14 | 16 |

s[]: study
a[]: study arm
time: follow-up time corresponding to $u_{m+\frac{1}{3}}$ . time will be transformed depending on type of model, e.g. for Weibull it will be log(time)
dt[,1]: time from begin of interval to $u_{m+\frac{1}{3}}$ ; dt[,2]: time from begin of interval to $u_{m+\frac{2}{3}}$ ; dt[,3]: time from begin of interval to $u_{m+1}$
r\_cond\_pfs1[]: observed number of patients who have not yet experienced progression or death at time $u_{m+\frac{1}{3}}$ ; n\_cond\_pfs1[] is the corresponding sample size at the beginning of the interval.
r\_cond\_pfs2[]: observed number of patients who have not yet experienced progression or death at time $u_{m+\frac{2}{3}}$ ; n\_cond\_pfs2[] is the corresponding sample size at the beginning of the interval.
r\_cond\_pfs3[]: observed number of patients who have not yet experienced progression or death at time $u_{m+1}$ ; n\_cond\_pfs3[] is the corresponding sample size at the beginning of the interval.
r\_cond\_os1[]: observed number of patients who have not died at time $u_{m+\frac{1}{3}}$ ; n\_cond\_pfs1[] is the corresponding sample size at the beginning of the interval.
r\_cond\_os2[]: observed number of patients who have not died at time $u_{m+\frac{2}{3}}$ ; n\_cond\_pfs2[] is the corresponding sample size at the beginning of the interval.

#### C.4 Simulation to assess performance of estimating stable-to-progression, stable-to-death, and progression-to-death hazard rates simultaneously based on conditional PFS and OS data for a given time interval

In order to assess the performance of estimating interval specific hazard rates for the stable-to-progression, stable-to-death, and progression-to-death transitions based on conditional PFS and OS data with the approach as proposed in this paper, we performed a simple simulation study for a single time interval. We set true values for each of the three transitions as follows: *{h*^*SP*^ = (0.1, 0.2, 0.3, 0.4, 0.5)*}, {h*^*SD*^ = (0.02, 0.05, 0.1)*}*, and *{h*^*SP*^ = (0.05, 0.1, 0.15, 0.2, 0.25)*}*. The sample size was assumed to be 1000. Based on the true rates and sample size, the number of PFS and OS events for four timepoints over a 3-month time interval were determined and subsequently transformed in three conditional PFS events and three conditional OS events according the approach described in the Appendix. This data was used to estimate *h*^*SP*^, *h*^*SD*^, and *h*^*P D*^ according to Equation 3, Equation 4, and Equation 5 for each simulation run. One hundred simulation runs were performed for each scenario.

Results are presented in Table A3, Figure A14, Figure A15, and Figure A16. The mean of the estimates for *h*^*SP*^, *h*^*SD*^, and *h*^*P D*^ across the hundred simulaton runs are close to the true hazard rates. The mean of the standard deviation of the posterior distribution for *log*(*h*^*SP*^) (equivalent to the mean of the standard error of the estimate with a frequentist analysis) across the hundred simulaton runs and the standard deviation of the estimates for *log*(*h*^*SP*^) across the same simulation runs were relatively similar across the different scenarios. However, for *h*^*SD*^, and expecially for *h*^*P D*^, the mean standard deviation of the posterior distribution is larger than the standard deviation of the estimates indicating that the obtained 95% credible intervals for these transitions with the proposed approach seem conservative. The proportion of 95% credible intervals that include the true hazard rate over the hundred simulation-runs is about 95% for *h*^*SP*^ but may be higher or less than 90% depending on the true rate of each of the three transitions as depicted in Figure A14. For *h*^*SD*^ and *h*^*P D*^, the proportion of 95% credible intervals that include the true hazard rates over the hundred simulation runs seems greater than 95%, indiciting some degree of overcoverage.

**Table A3:**
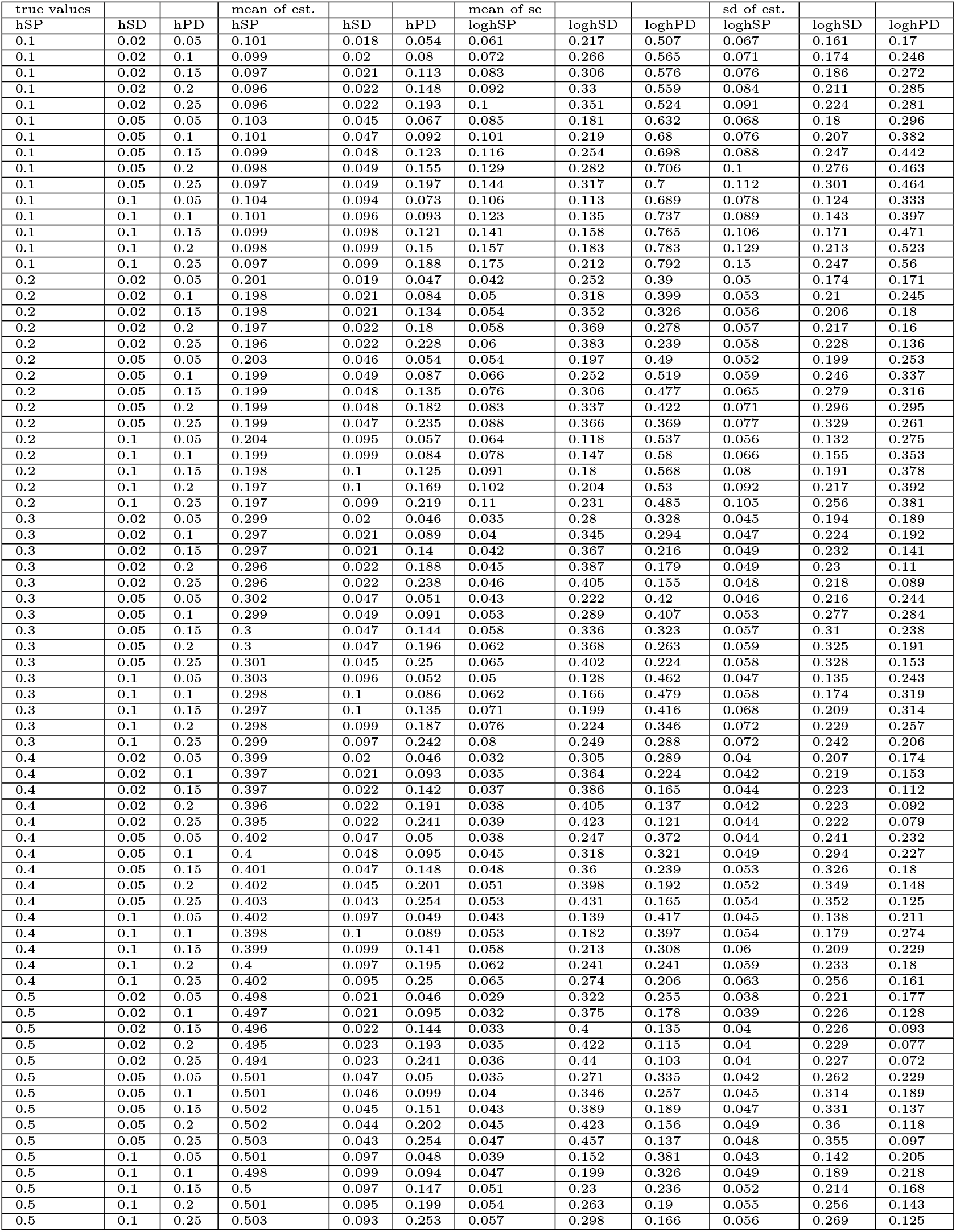
Performance of estimating simultaneously hazard rates for stable-to-progression, stable-to-death, and progression-to-death transitions based on conditional PFS and OS data for a given time-interval.

**Figure A14:**
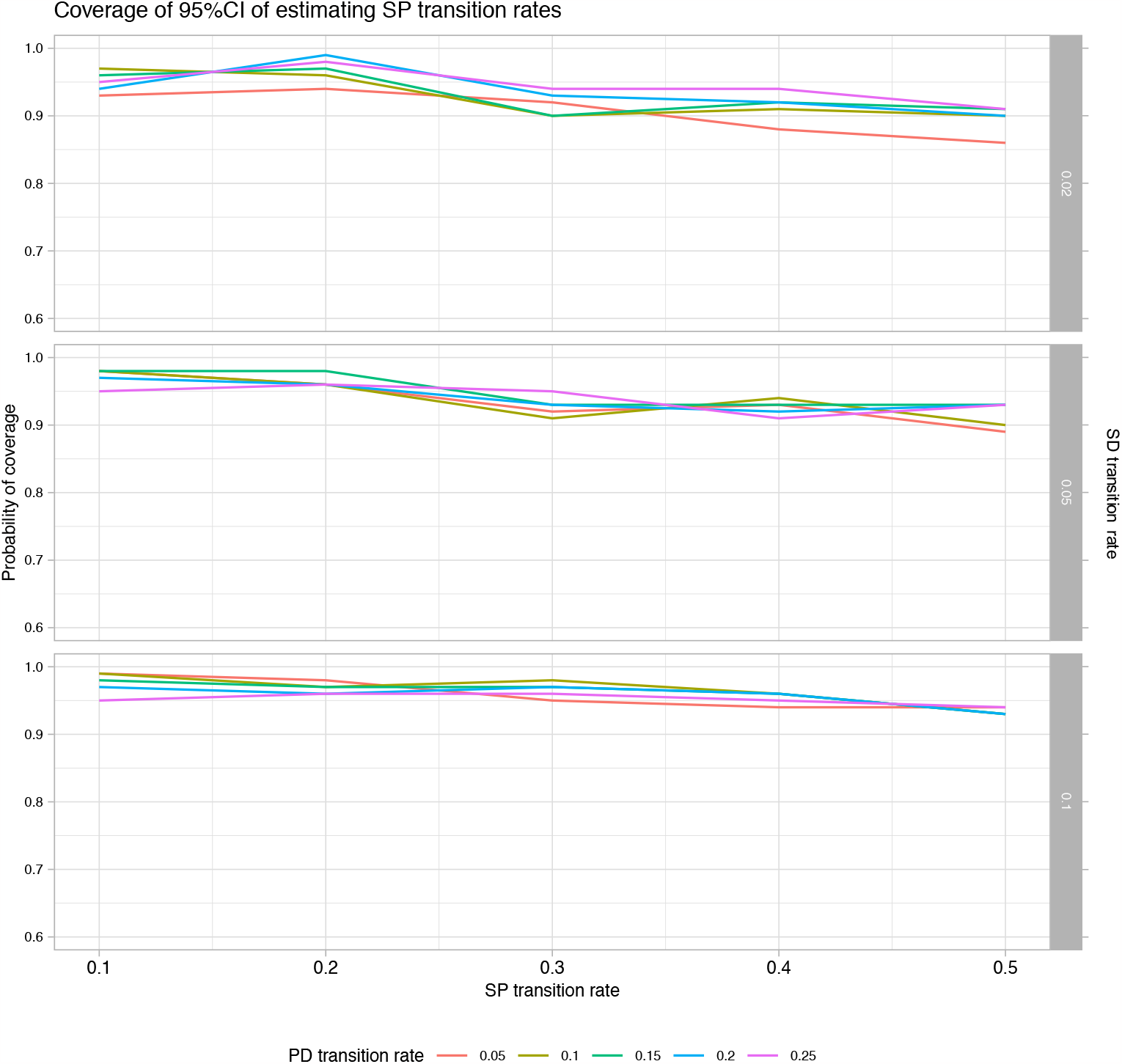
Coverage probablity of the 95% credible interval of the stable-to-progression hazard rate stratified by progression-to-death and stable-to-death rates.

**Figure A15:**
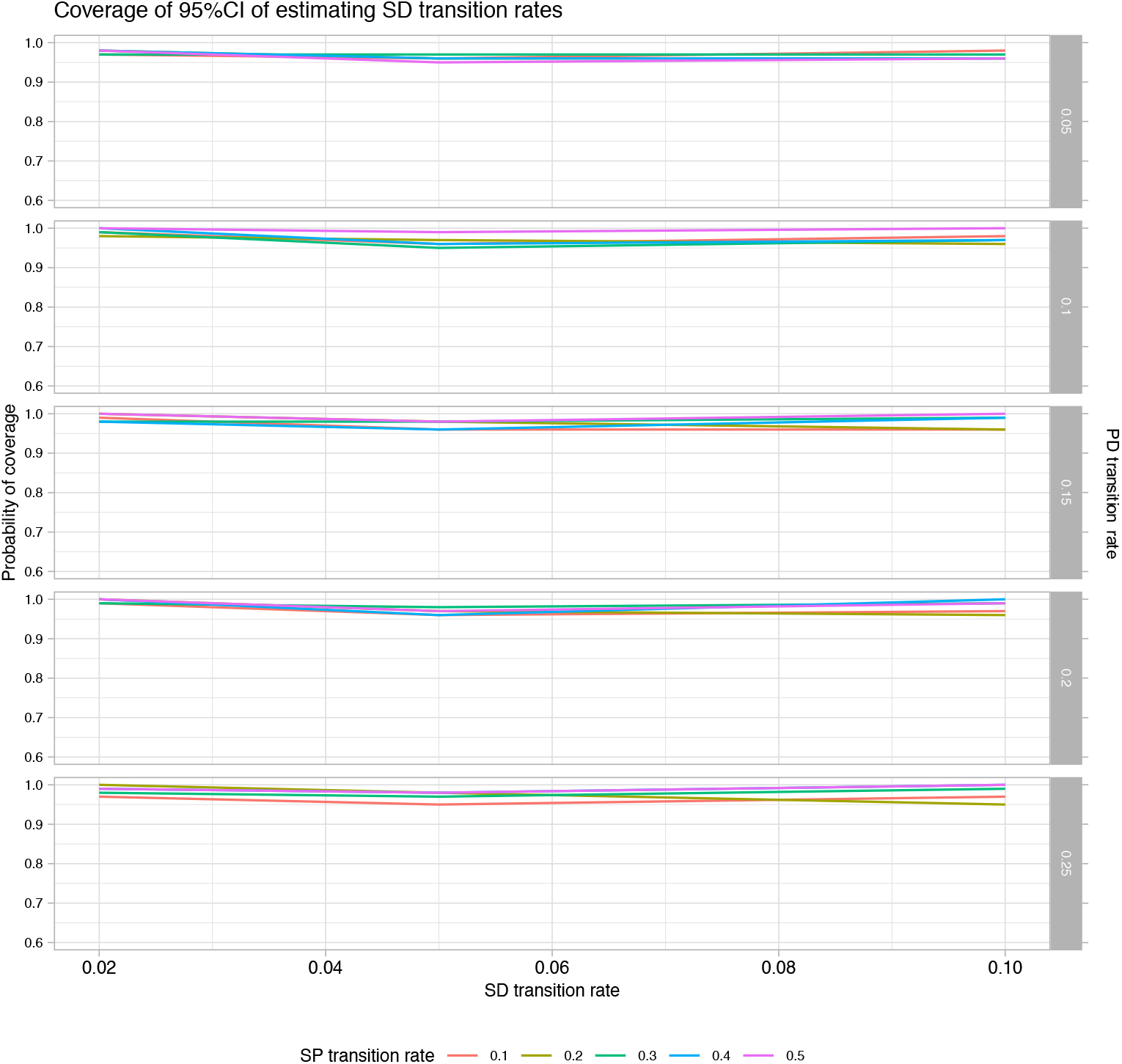
Coverage probablity of the 95% credible interval of the stable-to-death hazard rate stratified by stable-to-progression and progression-to-death rates.

**Figure A16:**
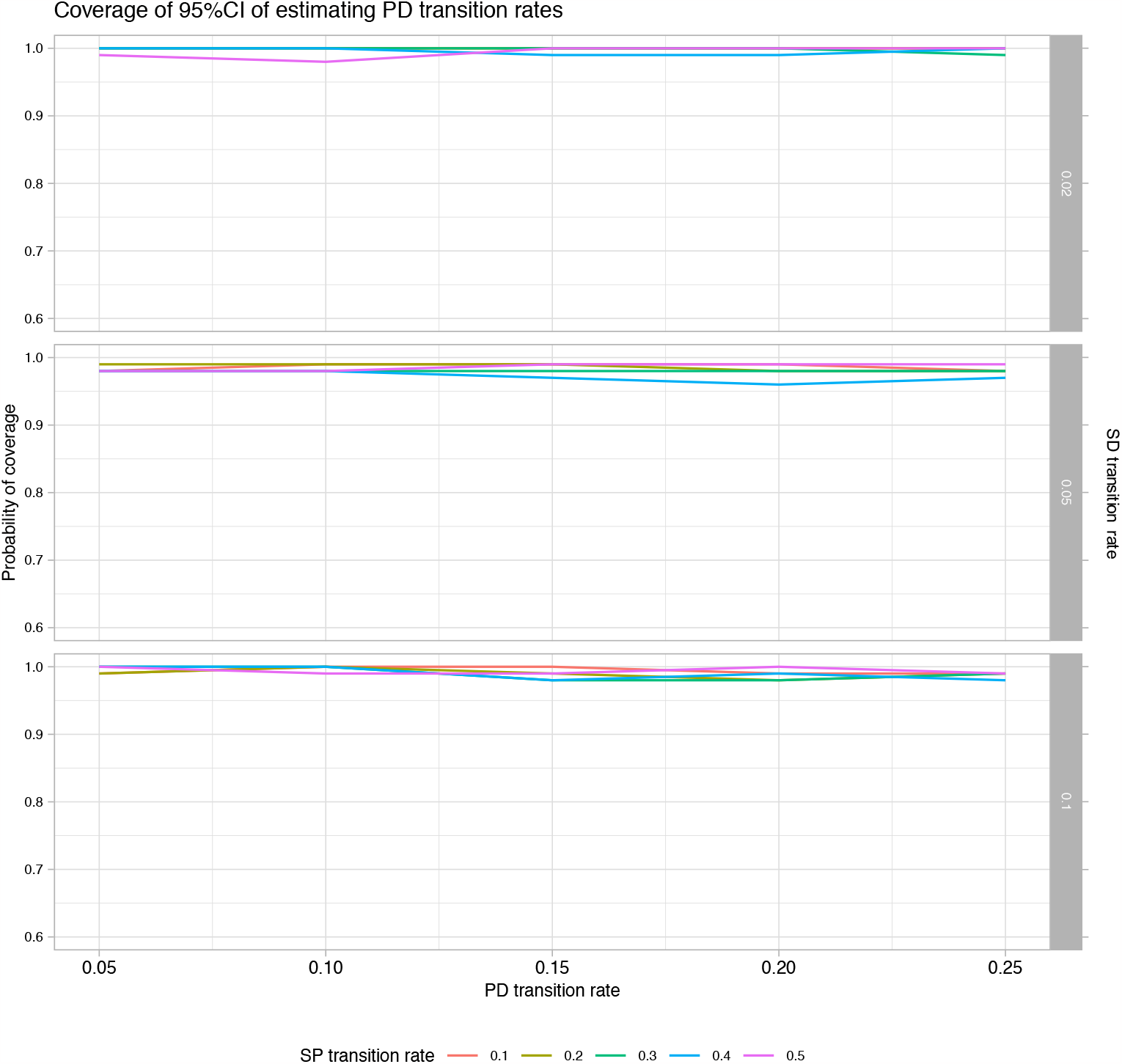
Coverage probablity of the 95% credible interval of the progression-to-death hazard rate stratified by stable-to-progression and stable-to-death rates.

#### C.5 Length of intervals to estimate hazards

In order to estimate the three parameters 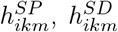, and 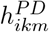 for each interval *m*, we defined Equation 3, Equation 4 and Equation 5 for three time points per 3-month interval: 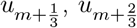, and 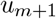. To assess the impact of the length of the interval on model estimates, we also estimated time-varying hazards for the different transitions as obtained with the meta-analysis of treatment 1 according to Model 1 based on an interval length of 1.5 months also using three data points. Out of the presented models for the example, Model 1 is the most flexible in terms of capturing changes in the hazards over time and therefore best suited to compare the impact of the length of the time-interval. The estimated hazards and patterns are similar between the two analyses, indicating that the method seems robust to the chosen interval length, at least for intervals between 1.5 and 3 months.

**Figure A17:**
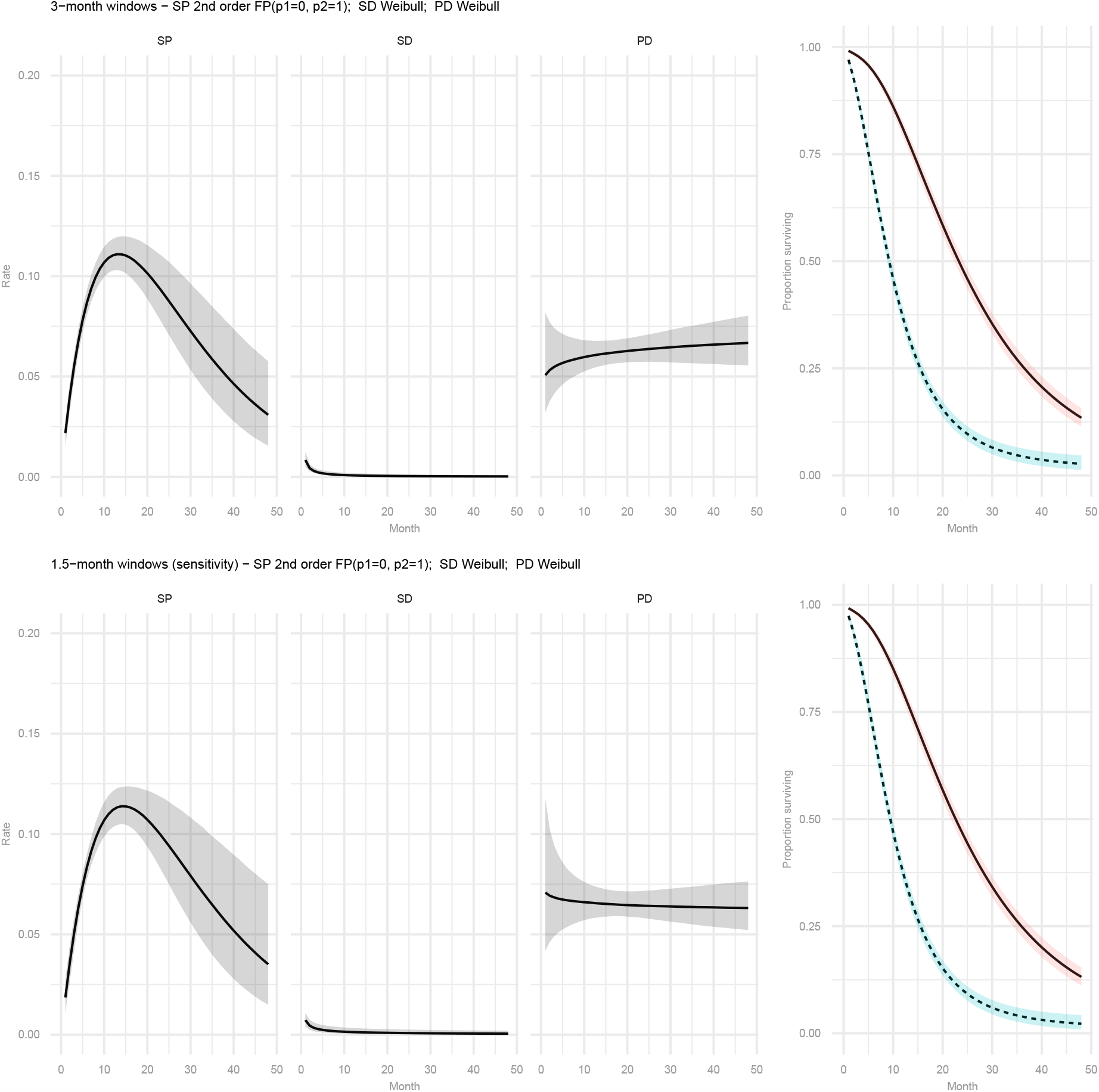
**Pooled estimates of hazard rates over time for the stable-to-progression transition (SP), stable-to-death transition (SD), and progression-to-death transition (PD), and PFS and OS curves with treatment 1 from a multi-state fixed effects meta-analysis model based on a 3-month interval or a 1.5-month interval**

## Notes

### Competing Interest Statement

The authors have declared no competing interest.

### Summary of Updates

Modified equation 2 to reflect the general RE model. Added a comparison of a 3-month vs 1.5-month window constant hazards in terms of impact on results. Revised paragraph 4 and 5 of discussion section. Added example dataset.

